# Characterising within-hospital SARS-CoV-2 transmission events: a retrospective analysis integrating epidemiological and viral genomic data from a UK tertiary care setting across two pandemic waves

**DOI:** 10.1101/2021.07.15.21260537

**Authors:** Benjamin B. Lindsey, Ch. Julián Villabona-Arenas, Finlay Campbell, Alexander J. Keeley, Matthew D. Parker, Dhruv R Shah, Helena Parsons, Peijun Zhang, Nishchay Kakkar, Marta Gallis, Benjamin H. Foulkes, Paige Wolverson, Stavroula F Louka, Stella Christou, Amy State, Katie Johnson, Mohammad Raza, Sharon Hsu, Thibaut Jombart, Anne Cori, Sheffield COVID-19 Genomics Group, The COVID-19 Genomics UK (COG-UK) consortium, CMMID COVID-19 working group, Cariad M. Evans, David G. Partridge, Katherine E. Atkins, Stéphane Hué, Thushan I. de Silva

## Abstract

**Objectives:** To characterise within-hospital SARS-CoV-2 transmission across two waves of the COVID-19 pandemic.

**Design:** A retrospective Bayesian modelling study to reconstruct transmission chains amongst 2181 patients and healthcare workers using combined viral genomic and epidemiological data.

**Setting:** A large UK NHS Trust with over 1400 beds and employing approximately 17,000 staff.

**Participants:** 780 patients and 522 staff testing SARS-CoV-2 positive between 1st March 2020 and 25th July 2020 (Wave 1); and 580 patients and 299 staff testing SARS-CoV-2 positive between 30th November 2020 and 24th January 2021 (Wave 2).

**Main outcome measures:** Transmission pairs including who-infected-whom; location of transmission events in hospital; number of secondary cases from each individual, including differences in onward transmission from community and hospital onset patient cases.

**Results:** Staff-to-staff transmission was estimated to be the most frequent transmission type during Wave 1 (31.6% of observed hospital-acquired infections; 95% CI 26.9 to 35.8%), decreasing to 12.9% (95% CI 9.5 to 15.9%) in Wave 2. Patient-to-patient transmissions increased from 27.1% in Wave 1 (95% CI 23.3 to 31.4%) to 52.1% (95% CI 48.0 to 57.1%) in Wave 2, to become the predominant transmission type. Over 50% of hospital-acquired infections were concentrated in 8/120 locations in Wave 1 and 10/93 locations in Wave 2. Approximately 40% to 50% of hospital-onset patient cases resulted in onward transmission compared to less than 4% of definite community-acquired cases.

**Conclusions:** Prevention and control measures that evolved during the COVID-19 pandemic may have had a significant impact on reducing infections between healthcare workers, but were insufficient during the second wave to prevent a high number of patient-to-patient transmissions. As hospital-acquired cases appeared to drive most onward transmissions, more frequent and rapid identification and isolation of these cases will be required to break hospital transmission chains in subsequent pandemic waves.

## Introduction

Severe acute respiratory syndrome coronavirus *2* (SARS-CoV-2) has resulted in multiple hospital outbreaks, exposing healthcare workers (HCWs) and non-COVID-19 patients to SARS-CoV-2 infection.[1–3] At least 32,307 patients are thought to have been infected in hospitals in the UK during the pandemic so far and an estimated 414 HCWs have died.[4] To safely continue routine and elective activities in hospitals during times of high SARS-CoV-2 incidence, it is important to discern factors that drive hospital-acquired infections. This greater understanding can be used to protect staff and patients, as well as informing further efforts to contain hospital outbreaks.

Identifying within-hospital SARS-CoV-2 transmission events using epidemiological data remains challenging for two reasons. Firstly, the high variability in the viral incubation period means it is often difficult to determine whether hospital onset cases are community- or hospital-acquired. Secondly, at least 33% of SARS-CoV-2 infections in adults are thought to be asymptomatic[5], therefore identifying all patients and HCWs contributing to transmission is challenging. In the limited instances where viral genomic data were analysed, this information was used to confirm or complement a purely epidemiological approach.[6–9] Elucidating the source of transmission events on the basis of viral genetic relatedness alone also entails considerable uncertainty due to the slow evolutionary rate of SARS-CoV-2.[10] In the time scale of an outbreak, a large proportion of individuals are infected by viruses too genetically similar to each other to distinguish genuine transmission events from unrelated infections. Furthermore, data from HCWs have rarely been included in previous analyses[11] and the relative role that patients and HCWs have played in fuelling hospital outbreaks in the UK remains largely unknown.

The integration of genomic, epidemiological and location data into a statistical inference framework offers a possible route to more accurate estimates of within-hospital transmission. Under such an approach, a transmission event between a pair of individuals is supported if their symptom onset times are compatible with the serial interval distribution SARS-CoV-2, if the individuals are in the same hospital location at the time of a suspected transmission event and if their viral genomes exhibit a high degree of relatedness.

In this study, we reconstructed SARS-CoV-2 outbreaks in a large NHS teaching hospital trust in England during the first two UK epidemic waves. We integrated over 2,000 viral genomic sequences, patient and staff locations, and routinely-available epidemiological information in a Bayesian framework that incorporates prior knowledge on the relative contributions of within-ward and between-ward transmission, as well as the proportion of unsampled cases.[12,13] Using this approach, we characterised the dynamics of SARS-CoV-2 transmission within a hospital setting, identifying key differences across the two pandemic waves, as well as the relative contribution of different groups and hospital locations to within-hospital transmission.

## Methods

### Study population

All cases in the study were patients or staff who tested positive for SARS-CoV-2 at Sheffield Teaching Hospitals NHS Foundation Trust (STHNFT), Sheffield, UK, between 1st March 2020 and 24th January 2021. STHNFT is a large UK NHS hospital Trust which includes five hospitals, has an average bed occupancy of 1400, and employs approximately 17,000 staff. SARS-CoV-2 nucleic acid amplification tests (NAAT) were performed on nose and/or throat swabs throughout the pandemic in line with contemporaneous UK Department of Health and Social Care guidance[14], using Hologic Panther or an in-house dual E/RdRp gene real time PCR assay.[15,16]

Patients were included in the analysis if they tested positive for SARS-CoV-2 at or during admission. Staff were included if they tested positive for SARS-CoV-2 and had worked in a clinical area in the 14 days prior to a positive test. Information on symptom onset of patients and their ward movements, together with place of work for staff, were extracted from STHNFT electronic records, when available.

### Sample Preparation, ARTIC Network PCR and Nanopore Sequencing

Sequencing was attempted on all available residual samples collected for routine diagnostic testing from STHNFT throughout the study period, with fluctuation in the proportion of positive samples sequenced due to multiple factors, including laboratory capacity and availability of stored samples. The first positive sample from each individual was selected for sequencing. RNA was extracted from viral transport medium and subject to the ARTIC network tiled amplicon protocol[17], followed by sequencing on an Oxford Nanopore GridION X5. Base calling was performed using a high accuracy model and the default basecaller in MinKNOW (currently guppy v4). Reads were filtered based on quality and length (400bp to 700bp) and mapped to the Wuhan reference genome (GenBank accession number NC_045512). Reads were downsampled to 200x coverage in each direction and variants called using nanopolish[18] to determine changes from the reference, followed by consensus sequence generation. Samples with over 90% genome coverage were included for further analysis. Viral genomic sequences were classified into PANGO lineages using the *Phylogenetic Assignment of Named Global Outbreak LINeages* (PANGOLIN)[19] version 2.4.2 and a multiple sequence alignment built using MAFFT[20] with 10 iterative refinements. All alignment positions flagged as problematic for phylogenetic inference were removed, including highly homoplasic positions and 3′ and 5′ ends [21].

### Hospital outbreak reconstruction model

To identify likely transmission events between individuals, we extended the most recent implementation of the *outbreaker2* model to capture ward-level transmission[12,13]. Our Bayesian model calculates the likelihood of a transmission event from case *i* to case *j* at a putative transmission time given the time of symptom onset for case *i* and *j*, the Hamming distance between the corresponding virus genetic sequences, and the ward that *i* and *j* were on at the time of infection. To formulate the likelihood, we used information on the generation time distribution (the delay between the infection of a primary case and the infection of a secondary case), the incubation period distribution (the delay between the infection and symptom onset) and the assumption that an infection happened within a ward rather than between individuals on different wards.[22,23] We defined the ascertainment probability as the proportion of cases that were captured in our dataset and calculated this as the product of (i) the proportion of known cases with high quality sequence, (ii) the proportion of these cases where the ward location was known, and (iii) the proportion of all cases that were likely detected before discharge via testing. In our base case analysis, we used a global sensitivity method to capture uncertainty in (i) the proportion of cases that were imported community-acquired infections, and (ii) the symptom onset date for individuals for whom this information was unavailable and (iii) multiple work locations for some staff. We also used a point estimate for our ascertainment probability but varied this estimate in a one-way sensitivity analysis (**Full details in File S2**). A final posterior distribution of 10,000 transmission networks was inferred by integrating over the uncertainty across imputed datasets of symptom onset times, multiple staff work locations and the number (and identity) of community-acquired cases. Full details of the model, including model fitting and prior distributions, are provided in **Table S1**. All code is available at https://github.com/Chjulian/sheffield_HT.

### Ethical approval

Approval for the study was obtained from the UK Health Research Authority (IRAS 281918), with sequencing performed according to The COVID-19 Genomics UK (COG-UK) study protocol approved by the Public Health England Research Ethics Governance Group (R&D NR0195).

## Results

### Study population, SARS-CoV-2 testing and infection control measures

During the first wave of the UK epidemic (Wave 1; defined as 1st March to 25th July 2020 for our analysis), 886/1,184 (74.8%) patients and 842/1,104 (76.3%) HCWs at STHNFT who tested positive for SARS-CoV-2 had sequence data available with over 90% genome coverage (**Figure 1**). During the second wave (Wave 2; defined here as 30th November 2020 to 24th January 2021) 669/1,183 (56.6%) SARS-CoV-2 positive patients and 651/838 (77.7%) SARS-CoV-2 positive HCWs had sequence data available with over 90% genome coverage. Cases were excluded if they were outpatients, non-clinical staff, non-STHNFT or community-based staff, household contacts of staff members, and staff who had missing ward location data, leaving 1,302 individuals in Wave 1 and 879 individuals in Wave 2 for the analysis (**Figure 1B, Table 1**).

**Table 1.** Summary of study cohort. Number of hospital locations includes wards and non-clinical areas. Ward movements refers to between ward movements and does not include bed movements within the same ward. Classification of patient cases according to likely source of infection (community or hospital-acquired) is based on SAGE criteria[25]. Community onset-community associated = positive test up to 14 days before or within 2 days after hospital admission; Community onset-suspected healthcare associated = positive test up to 14 days before or within 2 days after admission, with discharge from hospital within 14 days before test; Hospital onset-intermediate healthcare associated = positive test 3-7 days after hospital admission; Hospital onset-suspected healthcare associated = positive test 8-14 days after admission or 3-14 days after admission with discharge from hospital in 14 days before test; Hospital onset-healthcare associated = positive test 15 or more days after hospital admission.

|  | Wave 1 | Wave 2 |
| --- | --- | --- |
| Dates | 01/03/2020 to 25/07/2020 | 30/11/2020 to 24/01/2021 |
| Total cases | 1302 | 879 |
| Patients | 780 (59.9%) | 580 (66.0%) |
| Staff | 522 (40.1%) | 299 (34.0%) |
| Number of hospital locations | 120 | 93 |
| Global PANGO Lineages | 64 | 24 |
| Fraction of patients with inferred symptom onset date | 30.1% | 22.0% |
| Median ward movements per patient (range) | 3 (1-9) | 2 (1-9) |
| Patient classification |  |  |
| Community onset - community associated | 486 (62.3%*) | 236 (40.7%*) |
| Community onset - suspected healthcare associated | 79 (10.1%*) | 67 (11.6%*) |
| Hospital onset - healthcare associated | 80 (10.3%*) | 100 (17.2%*) |
| Hospital onset - suspected healthcare associated | 74 (9.5%*) | 88 (15.2%*) |
| Hospital onset - indeterminate healthcare associated | 61 (7.8%*) | 89 (15.3%*) |

\* Percentage of patient cases

**Figure 1.**
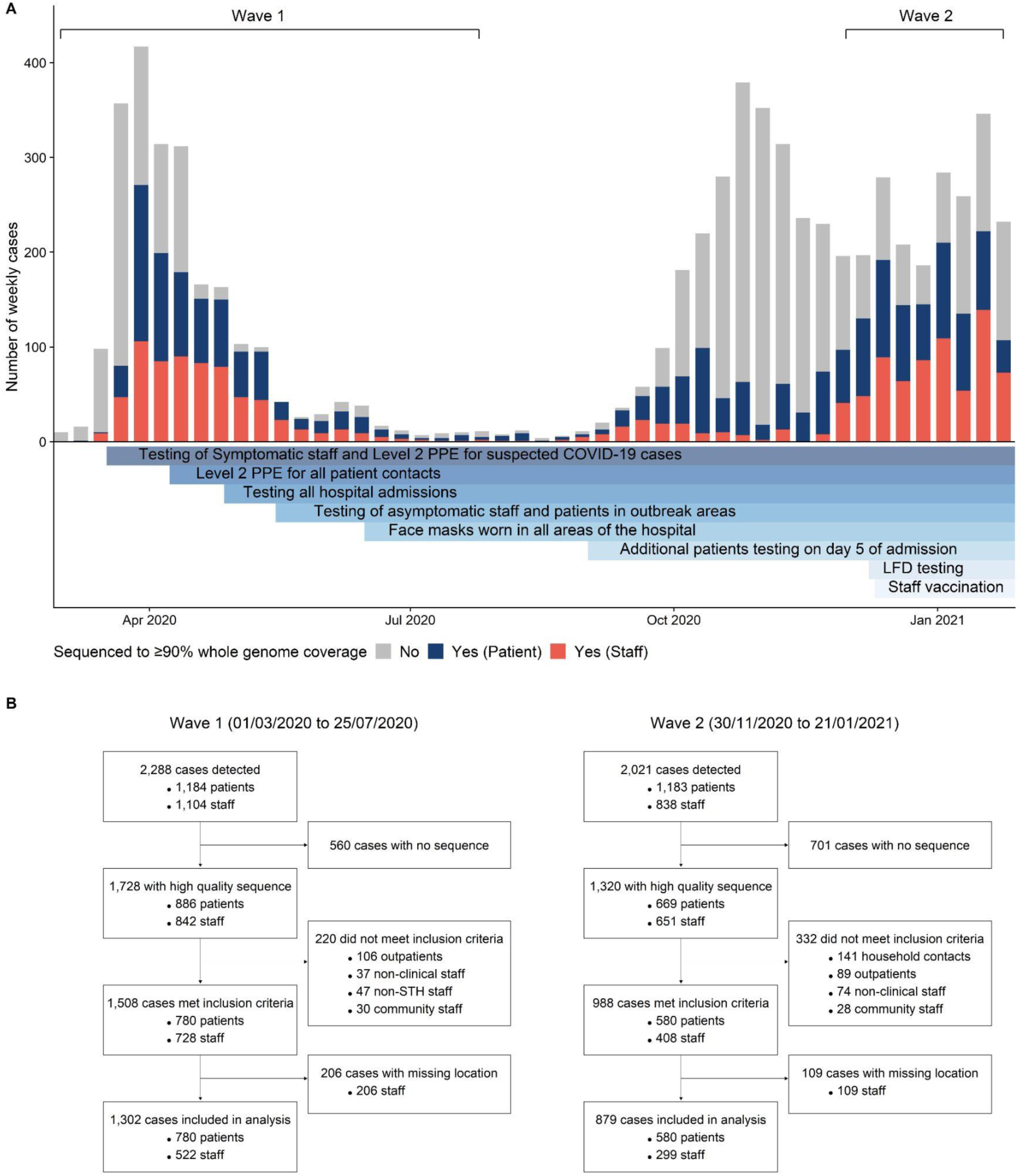
SARS-CoV-2 positive staff and healthcare worker samples included in the study. (A) SARS-CoV-2 positive cases in patients and staff at Sheffield Teaching Hospitals NHS Foundation Trust (STH) over time, and implementation of testing, prevention and control interventions. All SARS-CoV-2 nucleic acid amplification tests from STH patients and staff found to be positive in the hospital (pillar 1) diagnostic laboratory are shown in grey bars. Sequences with over 90% genome coverage for samples from healthcare workers (red) and patients (blue) are shown. These relate to samples with high quality sequences (1,728 in Wave 1 and 1,320 in Wave 2) shown in **Figure 1B**. Level 2 positive protective equipment (PPE) - aprons, gloves, eye protection and fluid resistant surgical face mask. LFD testing - Lateral flow device testing for staff two times per week. Grey bars represent SARS-CoV-2 cases tested (staff and patients) testing positive in the STH pillar 1 diagnostic laboratory with no high quality sequence available. Wave 1 and Wave 2 denote periods and samples included in the study. (B) Details of SARS-CoV-2 positive cases from patients and healthcare workers sequenced and included in the study.

SARS-CoV-2 testing policy and infection prevention and control (IPC) measures evolved throughout the pandemic (**Figure 1A**). Testing was performed in symptomatic patients with suspected SARS-CoV-2 infection on admission throughout the study period, with testing offered to symptomatic staff from 17^th^ March 2020.[24] Testing of all admissions regardless of symptoms commenced on 25^th^ April 2020 and screening of all asymptomatic patients and staff on wards with outbreaks from 18^th^ May 2020. In addition to screening on admission, all patients were routinely tested on day 5 of admission from 1^st^ Sep 2020. Routine twice weekly testing using lateral flow devices, followed by confirmatory NAAT, was offered to staff in all clinical areas from 8th December 2020. Level 2 personal protective equipment (PPE; aprons, gloves, eye protection and fluid resistant surgical face mask) was used by staff only for seeing suspected COVID-19 cases from 17^th^ March 2020, and for all patient contact from 8^th^ April 2020. HCWs were mandated to wear surgical face masks in all areas of the hospital from 15^th^ June 2020. The SARS-CoV-2 staff vaccination programme commenced on 10th December 2020.

Viral genomes were classified into 64 different PANGO lineages for Wave 1 and 24 lineages for Wave 2. Lineages B.1.1.1 (471/1,302, 36.2%), B.1.1.119 (180/1,302, 13.8%) and B.1 (110/1,302, 8.4%) predominated during Wave 1, while lineages B.1.177 (293/879, 33.3%), B.1.1.7 (263/879, 29.9%) and B.1.177.4 (112/879, 12.7%) predominated during Wave 2 (**Figure S1)**.

### Quantifying hospital-acquired infections

Using admission dates, symptom onset dates, SARS-CoV-2 positive test dates, ward location of cases and comparison between consensus viral genome sequences, our model inferred likely hospital transmissions between individuals, together with (i) the time of those events, (ii) the ward on which the transmission occurred, and (iii) whether the infector was a sampled or unsampled case.

Our model identified 473 (95% credible interval (CI) 310 to 688) hospital-acquired infections among our study population during Wave 1, of which 85 (95% CI 21 to 192) were unsampled. During Wave 2, there were 402 (95% CI 293 to 538) estimated hospital-acquired infections, of which 52 (95% CI 11 to 120) were unsampled. In Wave 1, patient cases comprised 40.9% (95% CI 36.1 to 45.5%) of all sampled hospital-acquired cases, compared to 65.1% (95% CI 60.3 to 69.4%) of all sampled hospital-acquired cases in Wave 2. Our model estimates showed good agreement with previous *a priori* inpatient epidemiological definitions of community or hospital onset categories (**Table 1 and Table S2**).[25] Specifically, no ‘community onset-community associated’ patient cases were identified as hospital-acquired by our model during either wave, and the majority of ‘hospital onset-hospital acquired’ and ‘hospital onset-suspected hospital acquired’ cases were identified as likely hospital-acquired by our model.

### Transmission chain reconstruction

We identified 95 (95% CI 82 to 109) transmission chains (defined as contiguous transmission events between 2 or more cases) in Wave 1 and 72 (95% CI to 61-84) transmission chains in Wave 2 (**Figure S2**). The median number of cases per transmission chain was 3 (95% CI 3 to 4) in Wave 1 and 4 (95% CI 3 to 5) in Wave 2. A staff member was identified as the index case in 50.6% (95% CI 42.0 to 58.0%) of transmission chains in Wave 1 and in 31.3% (95% CI 23.1 to 39.7%) of transmission chains in Wave 2. Forty different PANGO lineages were involved in transmission chains in Wave 1 and 13 were found in Wave 2 chains.

Of the transmissions between sampled cases in Wave 1, 31.6% (104/329, 95% CI 26.9 to 35.8%) were staff-to-staff events, 27.1% (89/329, 95% CI 23.3 to 31.4%) were patient-to-patient, 25.5% (84/329, 95% CI 22.1 to 29.3%) were patient-to-staff and 15.5% (51/329, 95% CI 12.2 to 19.1%) were staff-to-patient (**Figure 2C**). By contrast, during Wave 2, the majority of transmission events (162/311; 52.1%, 95% CI 48.0 to 57.1%) between sampled cases were patient-to-patient events, with 21.2% (66/311, 95% CI 18.0 to 24.1%) patient-to-staff, 13.5% (42/311, 95% CI 10.1 to 17.5%) staff-to-patient and 12.9% (40/311, 95% CI 9.5 to 15.9%) staff-to-staff transmission events (**Figure 2C**).

**Figure 2.**
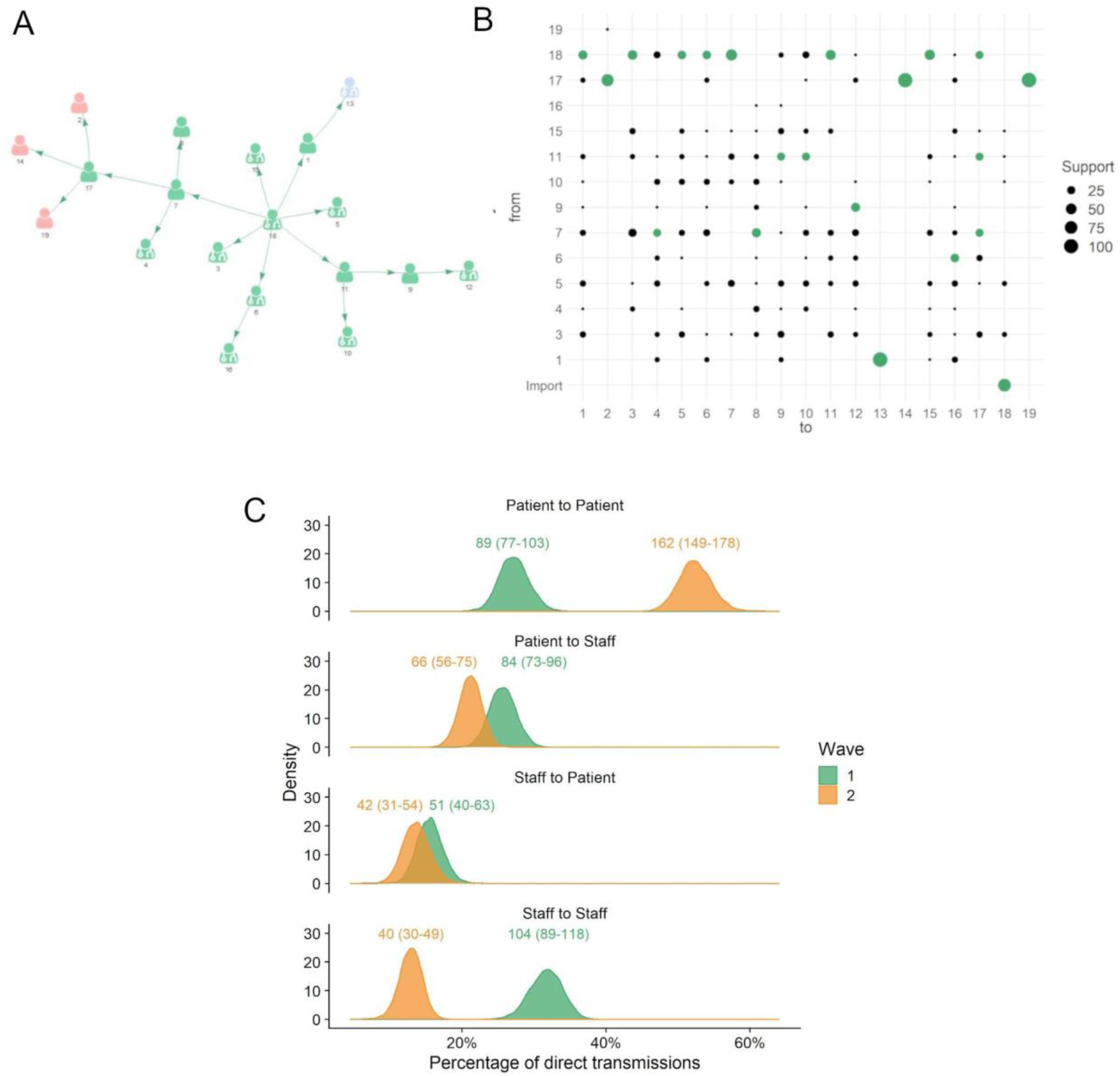
Within-hospital transmission chains and estimated infections within and between patients and healthcare workers. (A) An example transmission chain showing transmission between staff (icons with stethoscope) and patients (icons without stethoscope). Icons are labeled with their case identification number. Each color represents a separate ward where the infection occurred. (B) Uncertainty plot representing the support for each potential transmission pair shown in the example network in panel 2A. Support represents the percentage of networks across all those sampled, where a given infector is assigned to each case. Numbers on the plot correspond to the case identification numbers in Fig 2A and the green circles correspond to the transmission pairs displayed. Import = likely community-acquired infection imported into hospital. From = infector, to = infectee. (C) Comparison of the percentage of each transmission type between the two waves. The distributions display the percentages throughout the 10,000 plausible networks. The numbers above each distribution are the absolute numbers of each transmission pair with 95% credible intervals shown within the brackets.

In Wave 1, 55.3% (104/188, 95% CI 48.9 to 61.2%) of staff infections resulted from another staff case, which decreased to 37.7% (40/106, 95% CI 29.3 to 45.4%) in Wave 2. In Wave 1, 63.6% (89/140, 95% CI 56.1 to 71.0%) of patient infections resulted from another patient case which increased to 79.4% (162/204, 95% CI 73.7 to 84.6%) in Wave 2.

### Ward and Bay level transmission

Identified transmission events were not evenly distributed across the 132 hospital locations included, but isolated to 38 wards in 3 of the 5 hospitals within STHNFT. The 8 wards with the highest number of infections in Wave 1 accounted for 51.0% (95% CI 39.7 to 63.4%) of all transmissions, indicating the presence of transmission hot spots. A similar finding was observed during Wave 2, where 10 wards accounted for 50.1% (95% CI 40.6 to 60.5%) of all transmission events (**Figure 3**). We found evidence that the relative importance of specific wards in contributing to overall transmission was maintained across the two waves (Spearman’s Rank correlation Rho 0.54, *P*<0.0001, Ranked by mean number of transmissions per ward). However, there was considerable variability between waves, and several wards that were transmission hotspots in Wave 1 did not make up the 10 wards accounting for >50% of transmissions in Wave 2 (**Figure 3A-B**). Equally, several wards with no transmission event during Wave 1 were identified as transmission hotspots in Wave 2. Considerable variability was also seen between wards in the proportion of infectors and infectees made up by patients and staff (**Figure 3C-F**).

**Figure 3.**
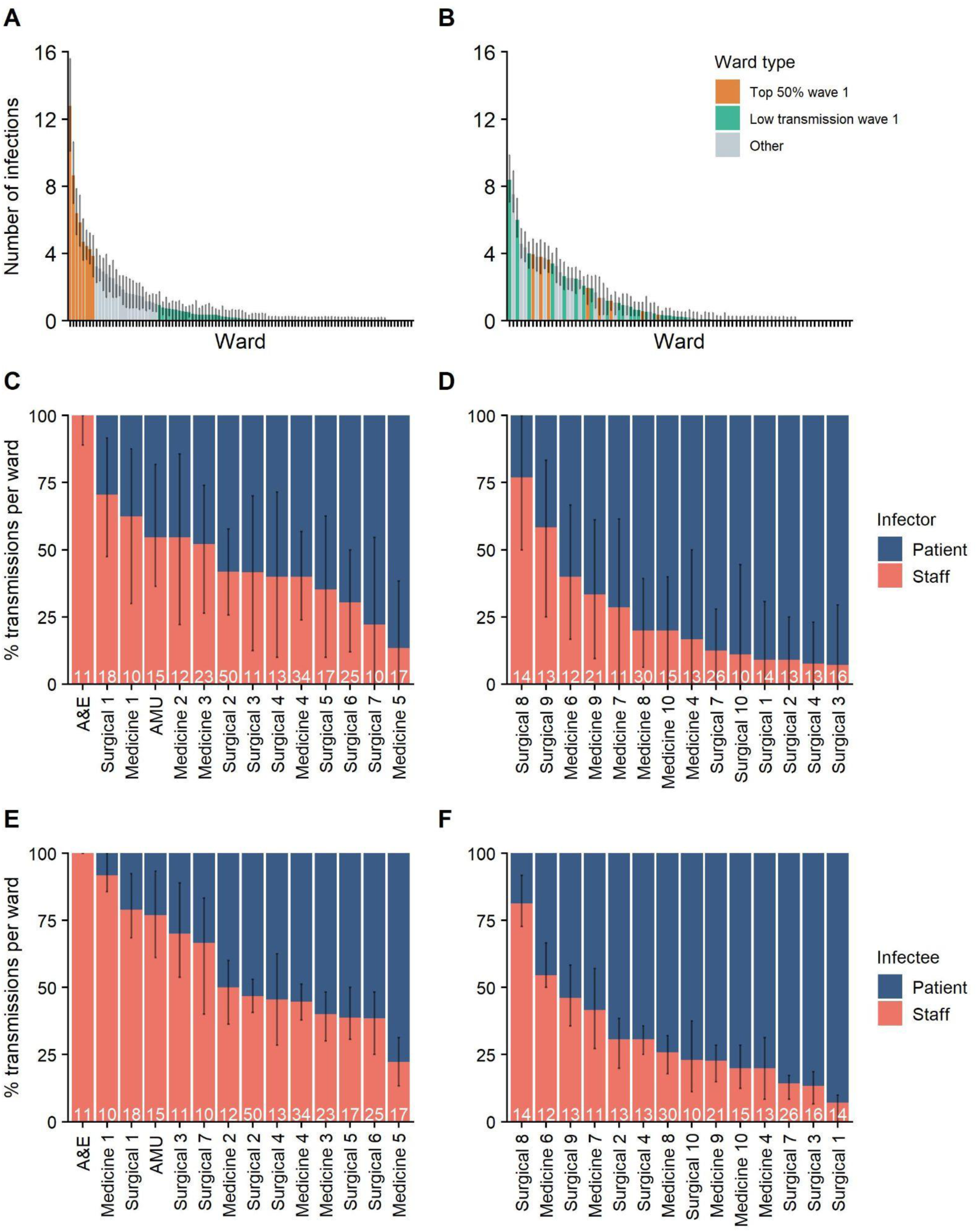
Hospital locations contributing to transmission events. Wards are ordered by the percentage of total transmission events identified in Wave 1 (A) and Wave 2 (B). Wards contributing to 50% of all transmission events in Wave 1 (n=8) are coloured in red in both Wave 1 and Wave 2 datasets. Wards with less than 1 transmission (mean of 10,000 networks) in Wave 1 are coloured in green in both Wave 1 and Wave 2 datasets. (C-F) Percentage of staff and patient infector and infectees per ward involved in transmission events. All wards with 10 or more transmissions in Wave 1 and Wave 2 are shown. Wards are ordered by the percentage of staff infector cases in Wave 1 (c) and Wave 2 (d) and percentage of staff infectee cases in Wave 1 (e) and Wave 2 (f). Numbers of transmission events per ward are shown at the bottom of each column. A&E = accident and emergency department; AMU = acute medical unit.

The highest number of infections on a single ward was 50 (95% CI 40 to 58) in Wave 1 but decreased to 30 (95% CI 23 to 36) in Wave 2. The highest number of separate transmission chains on a single ward was 8 (95% CI 5 to 11) for Wave 1 and 5 (95% CI 3 to 7) for Wave 2.

Wards comprise a combination of multi-bed bays with shared bathroom facilities and individual en-suite side rooms. We used a post hoc analysis to evaluate the contribution of bay-level transmission between patients to the outbreak. We identified 38.3% (95% CI 29.9 to 47.1%) of patient-patient transmissions in Wave 1 and 33.8% (95% CI 27.9 to 39.6%) in Wave 2 were between patients who shared a bay at some point during their stay. We estimated an increased risk of transmission between individuals who shared a bay compared with those who shared a ward as 2.8 (95% CI 2.2 to 3.5) times higher in Wave 1 and a 2.5 (95% CI 2.1 to 2.9) times higher in Wave 2.

### Secondary cases

The crude mean number of secondary cases was 0.30 (95% CI 0.21 to 0.38) for Wave 1 and 0.40 (95% CI 0.31 to 0.48) for Wave 2. Adjusting for unsampled cases, we estimated that 45.3% (95% CI 43.9 to 46.6%) of infections in Wave 1 and 43.6% (95% 42.3 to 45.0%) of infections in Wave 2 resulted in no onward transmission. Only 0.5% (95% CI 0.3 to 0.6%) infections in Wave 1 and 0.6% (95% CI 0.4 to 0.9%) infections in Wave 2 resulted in more than 5 secondary cases (**Figure 4**). Fewer patients classified as having community onset-community associated infections gave rise to secondary cases within the hospital (3.7%, 95% CI 1.8 to 5.6 for Wave 1; 3.5%, 95% CI 1.7 to 5.9 for Wave 2) compared to those with hospital onset-hospital acquired infections (45.5%, 95% CI 33.8 to 55.0 for Wave 1; 51.2%, 95% CI 41.0 to 60.0 for Wave 2), or other categories of hospital-onset cases (**Figure 4b, Table S2**). All findings were consistent across sensitivity analyses (**Figures S3-4**)

**Figure 4.**
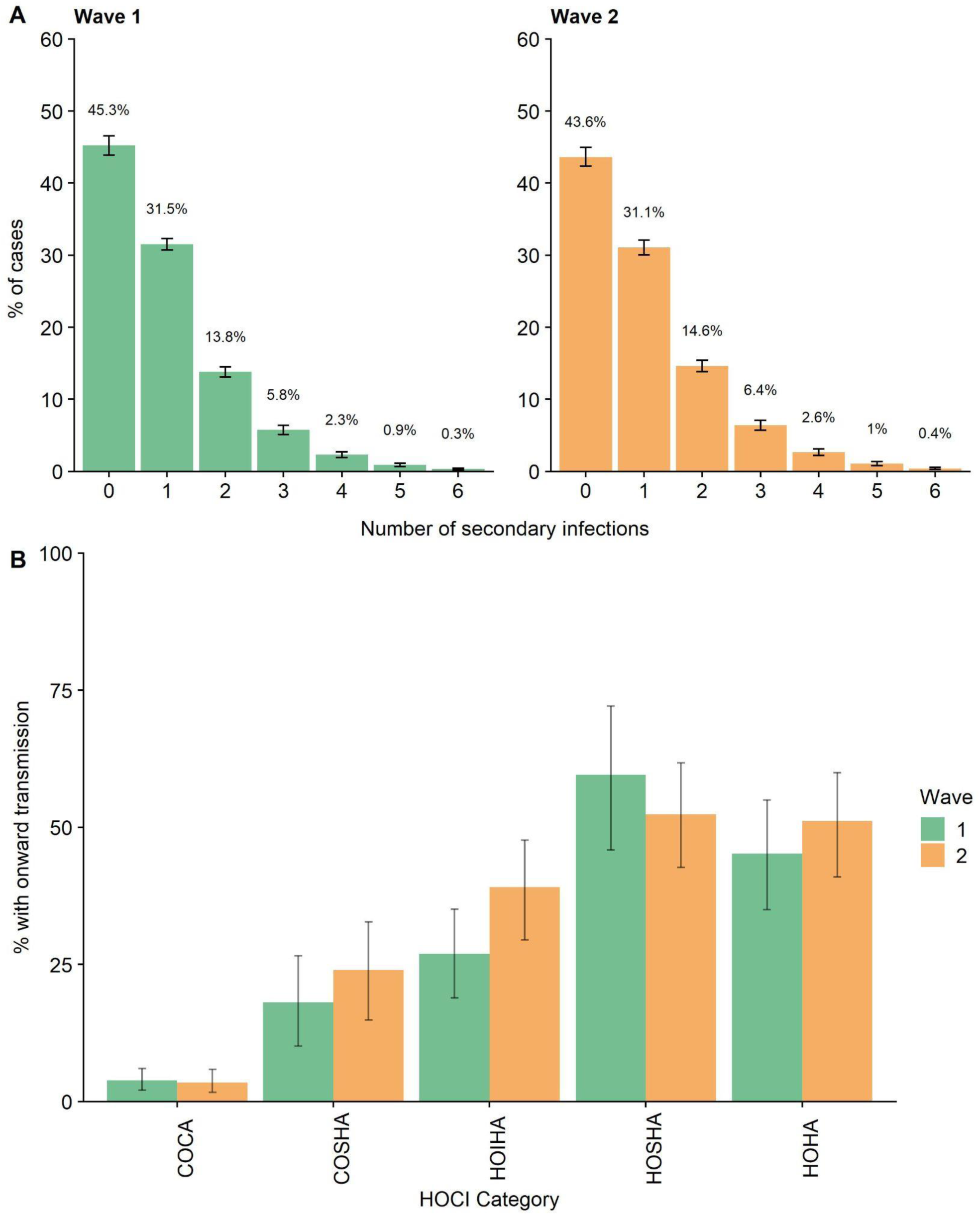
Secondary cases distributions and onward infections by HOCI category. (A) the percentage of cases with each number of secondary cases after adjusting for unsampled cases. (B) The percentage of each HOCI category case with at least 1 onward infection. COCA = Community onset community associated, COSHA = Community onset suspected hospital associated, HOIHA = Hospital onset indeterminate hospital associated, HOSHA = Hospital onset suspected hospital associated, HOHA = Hospital onset hospital associated. Classification of patient cases according to likely source of infection (community or hospital-acquired) is based on SAGE criteria[25]. Community onset-community associated = positive test up to 14 days before or within 2 days after hospital admission; Community onset-suspected healthcare associated = positive test up to 14 days before or within 2 days after admission, with discharge from hospital within 14 days before test; Hospital onset-intermediate healthcare associated = positive test 3-7 days after hospital admission; Hospital onset-suspected healthcare associated = positive test 8-14 days after admission or 3-14 days after admission with discharge from hospital in 14 days before test; Hospital onset-healthcare associated = positive test 15 or more days after hospital admission.

## Discussion

To our knowledge, our findings represent the largest collection of SARS-CoV-2 genomic and hospital epidemiology data to date used to reconstruct directional transmission networks, where we estimated hospital-acquired SARS-CoV-2 infections across two pandemic waves in the UK using a Bayesian framework. Importantly, our model accounts for unsampled events within these networks, which is crucial given the likely presence of unidentified infections or those lacking sequence data. We observed different contributions to the total number of within-hospital transmission events from those occurring between and within staff and patients across the two waves. We identified transmission hotspots within our institution, with a relatively small proportion of locations accounting for most hospital-acquired infections in staff and patients. We also found that the majority of SARS-CoV-2 infections resulted in onward transmission, with secondary cases identified in greater than 50% of infections but relatively few so called ‘superspreader’ events.

While much attention has been paid to staff potentially acquiring SARS-CoV-2 infections from patients due to perceived or real deficiencies in PPE, our findings suggest that the majority of HCW infections during the first pandemic wave were acquired from other HCWs. The contribution of these staff-to-staff infections to hospital-acquired transmission reduced dramatically during the autumn 2020 wave of SARS-CoV-2. Staff were less likely to initiate hospital transmission chains in Wave 2, accounting for 31.3% of index cases compared to 50.6% in Wave 1. Infection control practice and understanding of SARS-CoV-2 transmission evolved considerably during the pandemic. Improved social distancing and wearing of face coverings in non-clinical areas may explain some of these observations. In addition, the importance of asymptomatic transmission was increasingly appreciated and twice weekly lateral flow testing for healthcare workers was introduced in December 2020. Furthermore, seroprevalence rates of over 25% have been reported in HCWs following the first pandemic wave,[26] including in our NHS Trust,[27] which may have contributed to greater protection and reduced transmission in some areas. For example, we have reported that the SARS-CoV-2 seroprevalence in staff working on our acute medical unit by June 2020 was over 40%.[27] This was an area which was a hotspot for transmissions involving staff during our Wave 1 analysis but had very few transmission events identified during Wave 2. Nevertheless, by Wave 2, most staff infections in our NHS trust were estimated to have been acquired from patients, so further efforts are required to increase protection for HCWs. Staff vaccination is anticipated to have a large impact but is unlikely to have played a significant role in our observations due to introduction towards the end of Wave 2.

Hospital-acquired infections during Wave 2 were overwhelmingly dominated by patient-to-patient transmissions. The reasons for these events are likely to be multifactorial. UK hospitals faced significant bed pressures during this period and unlike during Wave 1, attempts were made to maintain as many routine and elective procedures for as long as possible. By this point, all patients in our hospitals were being routinely tested by NAAT for SARS-CoV-2 on admission and on day 5. Accordingly, the percentage of patients included in our dataset with asymptomatic infection increased from 10.4% during Wave 1 to 23.9% in Wave 2. The intense increase in patient-to-patient infections unfortunately occurred despite this enhanced focus on preventing asymptomatic transmission. Most of our transmission hotspots were wards built over two decades ago, with 6 – 8 beds per bay, and shared toilet facilities between every 1 to 2 bays.[28] While ventilation in these settings is in line with applicable regulations at the time of construction, none were designed with a respiratory pandemic in mind. Any contribution from these fixed estate issues will be challenging to address in a short timeframe. While viruses with greater transmissibility could also have played a role during Wave 2, circulation of the B.1.1.7/alpha variant occurred relatively late in our region compared to many other parts of the UK, and many Wave 2 transmission events were due to other SARS-CoV-2 lineages.

We found that the distribution of secondary cases was very similar across both waves, with over 50% of SARS-CoV-2 cases resulting in onward transmission, although only 10% of all infections resulted in more than two secondary cases. Our findings are different from those in a smaller study focusing on a few large clusters in another UK hospital, where 20% of individuals caused 80% of transmission events.[11] Importantly, we find that hospital-acquired SARS-CoV-2 cases give rise to a greater number of secondary cases than community-onset community-associated cases. Cases admitted from the community already suspected as having COVID-19 will have been isolated in single cubicles or COVID-19 cohort areas more rapidly, thus limiting opportunities for onward transmission. As severe disease requiring hospitalisation often occurs later in infection, they may also be at a less infectious stage, although hospitalised cases may also shed viable virus for longer.[29] In contrast, individuals with no SARS-CoV-2 symptoms who later acquired nosocomial infection may have initially be placed in bays with other susceptible patients, all of whom tested SARS-CoV-2 negative on screening tests at admission. Given the high viral loads during the first few days of infection, including during pre-symptomatic stages,[30] our data suggest that these individuals may acquire SARS-CoV-2 in hospital and have ample opportunity for onward transmission before being detected and isolated. This finding indicates that asymptomatic testing of patients on admission and day 5 was insufficient to prevent these scenarios. Daily testing of patients in the first week of admission or more regular testing throughout admission may allow greater opportunity for intervention, as well as more recent recommendations to IPC guidance such as routine wearing of masks by all patients in bays. Equally, rapid point-of-care testing (POCT) on admission may also reduce the window for transmission early in admission as it allows earlier isolation of asymptomatic community-acquired cases. Our Trust instituted POCT for all medical admissions in mid-January 2021.

Our study has several limitations that are important to consider. Firstly, despite the large number of individuals included, this is a single centre study and may not be generalisable across all UK hospitals given the heterogeneity in practice, building infrastructure, and patient population that exists. Our organisation had a high number of documented hospital-acquired infections in patients between March 2020 and March 2021 (n=795), but was not an outlier with 7 other NHS Trusts with higher numbers (highest n=1,463).[31] Seven of the top 10 busiest NHS Trusts (including our own) were also in the top 10 Trusts with the highest number of hospital-acquired COVID-19 infections in patients, indicating a common theme that may be a driver of nosocomial SARS-CoV-2 infections.[32] The effectiveness of various infection control measures on within-hospital transmissions over time in our setting is also likely to be generalisable to many UK hospitals, as they were based on national guidance applicable to all NHS Trusts. Many institutions will share similar issues regarding outdated infrastructure with 43% of NHS Trusts occupying estates greater than 30 years old.[33]

Although we did not have a selective sampling strategy, either for case detection or sequencing of positive cases, it is possible that there was an unobserved sampling bias. For example, as individuals with higher viral loads will be more infectious and their samples more likely to result in successful sequencing, they are more likely to have been included in our dataset. Our model attempted to account for unsampled events but would not mitigate any bias entirely. While we had electronic records of precise location data for patients during all times of their admission, staff location data were less granular and dependent on self-reported areas of work in the 14 days prior to infection. We also had to make several assumptions regarding priors in our model, but have attempted to undertake sensitivity analyses to test any potential impact of these.

With this study, we provide evidence that the integration of clinical surveillance data, viral genomic information and modelling enhances our capacity to unravel the complex transmission dynamics of SARS-CoV-2 in times and places of high incidence. The application of such a high-resolution framework to healthcare settings offers attractive perspectives for guiding the development of a safe environment for both staff and patients, as it may have a significant impact on the reduction of SARS-CoV-2 hospital transmission in subsequent epidemic waves.

## Contributions

Contributor roles were assigned as per http://credit.niso.org/. BBL, CJVA, FC, KEA, SH & TIdS were involved in the conceptualisation of the study. BBL, AJK, MDP, DRS, PZ, NK, MG, BHF, PW, SFL, SC, AS, KJ, MR, SH, HP, CME, DGP, KEA, SH and TIdS were involved in data collection and curation. BBL, CJVA, FC, MDP, SH and KEA were involved in data analysis. AC, TJ, SH, KEA and TIdS were involved in supervision of the project. BBL and CJVA were involved in data visualisation. FC was involved in software development. BBL, CJVA, FC, KEA, SH & TIdS wrote the original draft. All authors were involved in reviewing and editing the final manuscript.

## Data Availability

Data is available upon request however a data sharing agreement is required

## Acknowledgements

We thank the Sheffield Bioinformatics Core for their thoughtful discussion. We would like to thank the members of the Sheffield Biomedical Research centre for their continued support of the SARS-CoV-2 sequencing work in Sheffield. We thank all partners of and contributors to the COG-UK consortium, who are listed at https://www.cogconsortium.uk/about/. Sequencing of SARS-CoV-2 samples was undertaken by the Sheffield COVID-19 Genomics Group as part of the COG-UK CONSORTIUM and supported by funding from the Medical Research Council (MRC) part of UK Research & Innovation (UKRI), the National Institute of Health Research (NIHR) and Genome Research Limited, operating as the Wellcome Sanger Institute. MDP and DW are funded by the NIHR Sheffield Biomedical Research Centre (BRC - IS-BRC-1215-20017). TIdS is supported by a Wellcome Trust Intermediate Clinical Fellowship (110058/Z/15/Z). CJVA and KEA were funded by an ERC Starting Grant (action number 757688). This study is partially funded by the NIHR Health Protection Research Unit in Modelling and Health Economics, a partnership between Public Health England, Imperial College London and LSHTM (grant code NIHR200908); and acknowledges funding from the MRC Centre for Global Infectious Disease Analysis (reference MR/R015600/1), jointly funded by the UK MRC and the UK Foreign, Commonwealth & Development Office (FCDO), under the MRC/FCDO Concordat agreement and is also part of the EDCTP2 programme supported by the European Union. *Disclaimer: “The views expressed are those of the author(s) and not necessarily those of the NIHR, Public Health England or the Department of Health and Social Care*.*”*

