## Supplementary appendix for "Characterising within-hospital SARS-CoV-2 transmission events: a retrospective analysis integrating epidemiological and viral genomic data from a UK tertiary care setting across two pandemic waves"

**Index**

**Supplementary File S1:** Outbreaker model specification **2**
Posterior distribution **2**
Ward likelihood **3**

**Supplementary File S2:** Supplementary methods **9**
Definitions of onset and association **9**
Ascertainment probability **9**
Symptom onset date imputation **11**
Proportion of imported cases **11**
Parameterisation of the importations **12**
Outbreaker model run specifications **14**
Secondary case distribution **14**

**Supplementary Figure S1.** Treemap of all PANGO lineages represented in the Wave 1 and Wave 2 datasets **17**

**Supplementary Figure S2.** Network of all transmission chains in Wave 1 and 2 **18**

**Supplementary Figure S3.** Sensitivity analysis of Wave 1 **19**

**Supplementary Figure S4.** Sensitivity analysis of wave 2 **20**

**Supplementary Figure S5.** Schematic representation of the outbreaker ward model **21**

**Supplementary Figure S6.** Integrating over transmission pathways to a given ward **22**

**Supplementary Figure S6.** Trace of the log-posterior values and in exploratory runs **23**

**Supplementary Table S1.** Notation of outbreaker2 model **24**

**Supplementary Table S2.** Percentage of each onset and association with infectors or onward secondary infections in the outbreaker model **25**

**Supplementary File S3:** Supplementary Authors **26**

**Supplementary File S4:** Additional References **47**

**Supplementary File S1: Outbreaker model specification**

**Posterior distribution**

The joint posterior distribution of the augmented data and parameters given the data can then be defined as:

$$P(A, \theta | D) \propto P(\theta) \prod_i \Omega_i^1 \Omega_i^2 \Omega_i^3 \Omega_i^4 \Omega_i^5 \Omega_i^6$$

where the likelihood terms model the incubation period ($\Omega_i^1$), generation time ($\Omega_i^2$), mutation events ($\Omega_i^3$), case reporting process ($\Omega_i^4$), contact process ($\Omega_i^5$) and ward infection process ($\Omega_i^6$). Given the absence of contact data, the contact term $\Omega_i^5$ is fixed at a value of 1.

The prior distributions of the parameters are assumed independent such that:

$$P(\theta) = p(\mu)p(\pi)p(\sigma)p(\tau)$$

The very high identity among SARS-COV-2 genetic sequences tends to link all the observations in the data and this results in posterior values for these parameters—for instance, $\pi$ converges to very high values indicating that the vast majority of the cases were present in our data—that are not compatible with clinical and epidemiological sources of information. To account for this (i) we assigned sensible fixed values to the ward parameters ($\sigma$ and $\tau$) and the ascertainment parameter ($\pi$) (**Supplementary Table S1**) and explored the variability around these values in one way sensitivity analysis; (ii) we modified the original outbreaker genetic likelihood by modelling the SNP distance between direct transmission pairs with a Poisson distribution; this places less likelihood weight on the genetic sequence data than the original formulation and allows a more evenly weighted integration with ward data. By modelling mutation events as a feature of infection, assuming no within-host diversity and using a per-infectious-generation, per-base mutation rate $\mu$ and number of comparable nucleotides $l(s_i, s_{\alpha_i})$, the genetic likelihood $\Omega_i^3$ of the SNP distance $d(s_i, s_{\alpha_i})$ between the samples of a case $i$ and its most recently sampled ancestor $\alpha_i$ separated by $\kappa_i$ generations is given by:

$$d(s_i, s_{\alpha_i}) \sim Poisson(\mu \times \kappa_i \times l(s_i, s_{\alpha_i}))$$

The mutation rate ($\mu$) represents a probability and was assigned a Beta distributed prior (**Supplementary Table S1**).

**Ward likelihood**

Though the infectious process is unobserved, the probability of infection between cases can be estimated from the ward data, more specifically from the pair of wards they are registered in on the most likely inferred day of infection (**Supplementary Figure S5**). We assume that infection is more likely to occur between individuals on the same ward, and model this assumption explicitly by defining the parameter $\sigma$ as the probability of a primary case infecting a secondary case registered on the same ward on the day of infection, conditional on the primary case having caused an infection. We refer to such events as "within-ward" infections.

The probability of a primary case infecting a secondary case registered on a different ward on the day of infection is therefore $1 - \sigma$, which we refer to as "between-ward" infections. This accounts for uncertainty in using ward data to estimate the infectious process, for example if a patient visits another ward during their stay in the hospital or if viral particles are transferred between wards via contaminated equipment or ventilation. However, $1 - \sigma$ defines the marginal probability of transmission pairs being registered on *any* two different wards, while the ward likelihood must consider the probability of infection occurring between the two specific wards where the transmission pair is based.

To calculate this probability, the likelihood of infection moving from one ward to another must be defined. Formally, given $N^w$ different wards in the hospital, we define a transition matrix $Q_{k,l}$ ($N^w \times N^w$) describing the probability of a case registered in ward $k$ infecting a case registered in ward $l$, conditional on a between-ward infection. The rows of this matrix therefore sum to 1, under the assumption that all outgoing infections occur in one of the other wards, and $Q_{k,k} = 0$. given that we lack information on the proximity between wards or the number of staff shared between them, we made the assumption that infection occurs with equal probability to any other ward such that $Q_{k,l} = 1/(N^w - 1)$.

Given the probability of within-ward infection $\sigma$ and the matrix of transition probabilities $Q_{k,l}$, we define a matrix X as a function of $\sigma$ as follows:

$$X(\sigma) = [\sigma\ (1-\sigma)Q_{1,2}\ \dots\ (1-\sigma)Q_{1,N^w}\ (1-\sigma)Q_{2,1}\ \sigma\ (1-\sigma)Q_{2,3}\ (1-\sigma)Q_{2,N^w}\ \dots\ (1-\sigma)Q_{3,2}\ \sigma\ \dots\ (1-\sigma)Q_{N^w,1}\ (1-\sigma)Q_{N^w,2}\ \dots\ \sigma\ ]$$

where individual elements $X(\sigma)[k,l]$ describe the probability of a case in ward $k$ infecting a case in ward $l$, conditional on the primary case having caused an infection). In other words, this matrix describes the expected proportion of infections originating in ward $k$ that end up in ward $l$. Using the ward designation of 0 to indicate a case not being registered in the hospital, we make the assumption that infection can only occur between cases in the hospital by defining $X(\sigma)[k, l] = 0$ if $k = 0$ or $l = 0$. This completes the ward likelihood for direct transmission pairs (**Supplementary Figure S5**), which describes the probability of a primary case on ward $k$ infecting a secondary case on ward $l$ on the day of infection of the secondary case, and is given by $X(\sigma)[k, l]$.

To keep the dimensionality of the ward model fixed, we analytically integrate over unobserved ward states and calculate the probability of the most recent sampled ancestor $\alpha_i$, registered on a given ward $k$, causing infection in case $i$, registered on a given ward $l$, after $\kappa_i$ generations of infection. This approach requires knowledge of the registered ward of $\alpha_i$ on the day of infection of the first unobserved case between $\alpha_i$ and $i$ (**Supplementary Figure S5**). However, the default *Outbreaker* model does not impute infection times of unobserved cases and as such this ward cannot be defined. We therefore impute an additional set of augmented data $T_i^{anc}$, called the ancestral infection time and defined as the infection time of the earliest unobserved case in an unobserved transmission chain separating $\alpha_i$ and $i$. $T_i^{anc}$ is thus only defined when $\kappa_i$ is greater than 1. A complete integration of $T_i^{anc}$ into the outbreaker likelihood would require calculating $P(T_i^{anc})$ as part of the generation time likelihood $\Omega_i^2$. However, this calculation would introduce variable model dimensionality that could only be explored using reversible-jump MCMC [1,2]. To avoid this complexity, we make the simplifying assumption that $P(T_i^{anc}) = 1$ for $T_{\alpha_i}^{inf} < T_i^{anc} < T_i^{inf}$ and $P(T_i^{anc}) = 0$ otherwise, and keep the original generation time likelihood $\Omega_i^2$.

Under these assumptions, the ward likelihood can be calculated by integrating over all potential transmission routes between the registered ward of $\alpha_i$ on day $T_i^{anc}$ and the registered ward of $i$ on day $T_i^{inf}$. For each unobserved case there exist two unobserved ward states, one on the day of infection and one on the day of onward infection (**Supplementary Figure S5**). As cases tend to remain on a single ward for a given period of time, these ward states cannot be considered independent. We therefore condition the later ward state on the earlier ward state by defining a parameter $\tau$ as the daily probability of a case being registered on the same ward it was in on the day of infection. If the case is found on a different ward, the specific probability of being transferred from a given ward $k$ to a different ward $l$ is calculated using the same transition probability $Q_{k,l}$ defined above. We therefore assume that ward transfers of unobserved cases follow the same transition probabilities as infections occurring between wards, though this assumption could be easily relaxed if the necessary data to inform these estimates were available. We then define a matrix Y as a function of $\tau$:

$$Y(\tau) = \left[\tau\ (1-\tau)Q_{1,2}\ \dots\ (1-\tau)Q_{1,N^w}\ (1-\tau)Q_{2,1}\ \tau\ (1-\tau)Q_{2,3}\ (1-\tau)Q_{2,N^w}\ \dots\ (1-\tau)Q_{3,2}\ \tau\ \dots\ (1-\tau)Q_{N^w,1}\ (1-\tau)Q_{N^w,2}\ \dots\ \tau\right]$$

where individual elements $Y(\tau)[k,l]$ indicate the probability of an unobserved case being transferred from a given ward $k$ on the day of infection to a given ward $l$ on the day of onward infection.

Given this parameterization it is possible to integrate over the unobserved transmission pathways linking a given ward $k$ to a given ward $l$ over $\kappa_i$ generations of infection. We do so by first defining all potential transmission routes between any two wards in a *single* unobserved generation of infection (**Supplementary Figure S6**). Fast integration over these transmission routes that accounts for all combinations of ward transfers and infection events can be achieved by matrix multiplication of $Y(\tau)$ and $X(\sigma)$ such that the elements of $Y(\tau) \times X(\sigma)$ describe the probability of an unobserved case that was infected on ward $k$ causing infection on ward $l$ after one unobserved generation of infection.

The probability of each route (**Supplementary Figure S6)** is conditional on the primary case becoming infected on that given ward in the previous generation of infection. The matrix product $Y(\tau) \times X(\sigma)$ can therefore be multiplied by itself in a recursive manner to integrate over transmission pathways for greater numbers of unobserved cases, where each multiplication by $Y(\tau) \times X(\sigma)$ represents the movement of infection in one unobserved generation.

This recursive approach can be initialised as the ward states of the most recent sampled ancestor $\alpha_i$ are observed. The probability of the first unobserved case becoming infected on any given ward $i$, given its infector $\alpha_i$ is registered in the observed ward $k$ on the day of onward infection, is simply given by $X(\sigma)[k,l]$. We therefore define a matrix Z as a function of $\sigma$, $\tau$ and $\kappa$:

$$Z(\sigma,\tau,\kappa_i) = X(\sigma) \times (Y(\tau) \times X(\sigma))^\wedge(\kappa_i - 1)$$

where × indicates the matrix multiplication operator and ^ the exponentiation operator. The first term calculates the probability of movement of infection from the sampled ancestor $\alpha_i$ to the first unobserved case, while the exponentiated term accounts for successive unobserved generations of infection. Individual elements $Z(\sigma, \tau, \kappa_i)[k, l]$ therefore describe the probability of infection originating in a given ward $k$ and ending up on ward $l$ after $\kappa_i$ generations of infection.

Given these probabilities, the ward likelihood $\Omega_i^6$ can then be defined for a given case $i$ as:

$$\Omega_i^6 = \{p\left(W_{i,T_i^{inf}} | \alpha_i, T_i^{inf}, W_{\alpha_i,T_i^{inf}}, \kappa_i, \sigma, \tau\right) \quad \kappa_i$$

$$= 1\ p\left(W_{i,T_i^{inf}} | \alpha_i, T_i^{inf}, T_i^{anc}, W_{\alpha_i,T_i^{anc}}, \kappa_i, \sigma, \tau\right) \quad \kappa_i > 1$$

and calculated as:

$$\Omega_i^6 = \{Z(\sigma, \tau, \kappa_i)\left[W_{\alpha_i,T_i^{inf}}; W_{i,T_i^{inf}}\right] \quad \kappa_i = 1\ Z(\sigma, \tau, \kappa_i)\left[W_{\alpha_i,T_i^{anc}}; W_{i,T_i^{inf}}\right] \quad \kappa_i > 1$$

The ward likelihood therefore describes the probability of infection originating in the ward that $\alpha_i$ is registered in on the day of onward infection ($T_i^{inf}$ if $\kappa_i = 1$ or $T_i^{anc}$ if $\kappa_i > 1$), and ending up in the ward that $i$ is registered in on its day of infection ($T_i^{inf}$) after $\kappa_i$ generations of infection.

**Supplementary File S2: Supplementary methods**

**Definitions of onset and association**

All COVID positive cases in patients were classified in 5 groups as in [4]:

(1) Community-Onset Community-Associated: Positive specimen date up to 14 days before, or within 2 days after, hospital admission, with no discharge from hospital in 14 days before specimen date.
(2) Community-Onset Suspected Healthcare-Associated: Positive specimen date up to 14 days before, or within 2 days after, hospital admission, with discharge from hospital in 14 days before specimen date.
(3) Hospital-Onset Suspected Healthcare-Associated: Positive specimen dated 8-14 days after hospital admission; or specimen date 3-14 days after admission, with discharge from hospital in 14 days before specimen date.
(4) Hospital-Onset Intermediate Healthcare-Associated: Positive specimen date 3-7 days after hospital admission, with no discharge from hospital in 14 days before specimen date.
(5) Hospital-Onset Healthcare-Associated: Positive specimen date to 15 or more days after hospital admission.

**Ascertainment probability $(\pi)$**

The ascertainment parameter, π, was estimated by combining (i) the proportion of known cases which have high quality sequence, (ii) the proportion of these cases where their location is known and (iii) the proportion of cases which are undetected. While the first two of these proportions were known, the third was more uncertain.

Wave 1 estimate

2,288 patient (1,184) and staff (1,104) cases tested SARS-CoV-2 positive at Sheffield Teaching Hospitals NHS Foundation Trust. 886/1184 (74.8%) patients and 842/1104 (76.3%) staff cases were sequenced to a genome coverage of greater than 90%. 220 cases were excluded because they were outpatients (106), non-clinical staff (37), external staff (47) or community based staff (30). 206 staff cases were excluded because their location of work was unknown. After exclusion, 86.4% (1,302/1,508) of staff and inpatient sequences were included. Combining the proportion of sequenced staff and inpatients known cases and included cases resulted in an ascertainment parameter of 65.4% (0.755*0.864*100). Assuming 80% of cases were detected, the reporting estimate for the model would be 52.3%.

Wave 2 estimate

2,021 patient (1183) and staff (838) cases tested positive during this time period. 669/1183 (56.6%) patient cases and 651/838 (77.7%) staff cases were sequenced to a genome coverage of greater than 90%. 332 cases were excluded because they were household contacts (141), non-clinical staff (74), community based staff (28) or outpatients (89). 109 staff cases were excluded because their location of work was unknown. After exclusion 89.0% (879/988) of sequenced staff and inpatient cases were included. Applying the same rationale as above results in a reporting parameter of 46.5%.

Based on these estimates, a sensitivity analysis was carried out varying the parameter from 20% to 60% for both waves.

**Symptom onset date imputation**

For 30.1% of wave 1 and 22.0% of wave 2 individuals, the symptom onset was not available. To account for these missing data, we estimated the distributions of delay (in days) between symptom onset and testing for inpatients and staff for whom this delay was known. For inpatients, we further stratified these distributions by six infection categories: Community Onset–Community Associated, Community Onset–Suspected Healthcare Associated, Hospital Onset–Healthcare Associated, Hospital Onset–Indeterminate Healthcare Associated, Hospital Onset–Suspected Healthcare Associated and undefined[3]. For staff, most were classified into a 'not admitted' category, with those who required admission stratified into the six categories listed above. All negative delays were removed from these 13 distributions. Next, for each individual for whom a date of onset was not recorded, we randomly sampled a delay between onset and testing from the respective known distributions associated with each infection category and individual type (staff or inpatient). If a distribution was unable to be defined due to absence of data, we instead sampled from the undefined patient or staff category. Finally, we calculated the imputed onset date of individuals with a sampled delay between symptom onset to testing by subtracting the sample delay from the recorded date of testing.

**Proportion of imported cases**

The precise proportion of imported cases was unknown and this uncertainty was captured as part of the final global sensitivity analysis. The proportion of imported patient cases was estimated from the admission date, symptom onset date and the incubation period. As we were unable to accurately estimate the proportion of imports for staff the sampling distributions outlined below include a staff import proportion ranging from 0% to 100%. The rationale for the Wave 1 and Wave 2 import proportions are outlined below.

<u>Wave 1 import proportion</u>

The estimated import proportion for patients in Wave 1 was 80% (~624/780; 95% confidence interval: 76% - 83%). Combining the inpatient and staff estimate (261/522, 50%) gives an estimated import percentage of ~68% (885/1302). Considering both extreme scenarios of no staff being imports and all staff being imports gives a range in the import percentage of ~48% to ~88%. To account for this uncertainty in the final results the import proportion for Wave 1 is varied 500 times, sampling from a normal distribution with mean 0.7 and standard deviation 0.075.

<u>Wave 2 import proportion</u>

Combining the patient (63%, ~365/580; 95% confidence interval: 58% - 67%) and staff (150/299, 50%) estimate gives an estimated import percentage of ~59% (515/879). Considering both extreme scenarios as above gives a range of ~42% to ~76%. The import proportion for wave 2 is sampled from the normal distribution with mean 0.6 and standard deviation 0.06.

**Parameterisation of the importations**

The *Outbreaker2* model requires the number of imports (i.e. community-associated infections) to be known in advance. Here, we relaxed this assumption by integrating over the distribution of the number of imports in independent MCMC chains, for each of which the number of imports was randomly drawn from a binomial distribution with N trials and a probability drawn from a Beta distribution (see proportion of imported cases section below). We define N as the total number of cases in the dataset, and the mean of the Beta distribution ($p$) as the probability across all cases that they are community-acquired. We first calculated, for each patient, the probability that they acquired the infection in the community as the probability that their incubation period was longer than their delay between admission and symptom onset. We then calculated the average patient-specific probability of a community-acquired infection. As admission data is unavailable for staff, we set bounds for the fraction of staff who acquired infection in the community as between 0 and 1. We then calculated the fraction of community-acquired infections across the whole dataset as the weighted sum of the patient-specific and the staff-specific probabilities as described in the proportion of imported cases section above.

To assign the identity of the imports, we calculated the mean individual likelihood $\Omega_i = \Omega_i^1 \Omega_i^2 \Omega_i^3 \Omega_i^4 \Omega_i^5 \Omega_i^6$ of case $i$ being infected by another individual observed in the dataset in a pre-run of the MCMC (1100 iterations with a thinning frequency of 1/50 and a burn-in of 500 iterations). For each MCMC chain in the main analysis, the randomly drawn number of imports were randomly assigned identities on the basis of the inverse of the individual likelihood $\Omega_i$.

**Outbreaker model run specifications**

Outbreaker was run in parallel integrating across uncertainty in the symptom onset data and the number and identity of imports. Specifically, we used a global sensitivity analysis in our base case analysis via an ensemble approach in which we generated a total of 10,000 posterior transmission networks. These 10,000 samples were generated from a sample of 100 imputed symptom onset datasets. In addition we drew five samples of the number and identity of imports and ran an MCMC across all 500 combinations. We then sampled 20 posterior networks from each of the converged chains of these 500 MCMC runs. Each parallel MCMC chain was run for 1100 iterations with a thinning frequency of 1/50 and a burn-in of 100 iterations. We ran the analysis using a timescale of quarter-day increments consistent with the ward location data being available to the nearest 6-hour interval.

**Secondary case distribution**

Our simulations calculate a posterior sample of the distribution of the number of observed secondary cases, X, by evaluating, for each case, the total number of connected cases (either direct transmission or with intermediate unsampled individuals). To calculate the true number of secondary cases (Z), including both observed and unobserved, we define the conditional probability,

$P(Z = j \mid X = i) = P(Y = j - i)$ for $j > i$

$P(Z = j \mid X = i) = 1 - \sum_{j>i} (1 - \pi)^{j-i}$ for $j = i$

$P(Z = j \mid X = i) = 0,$ else

We then calculate the unconditional probability distribution of true cases using the expression:

$$P(Z = j) = \alpha \sum_{i \leq j} P(Z = j \mid X = i) P(X = i)$$

where $\alpha$is the normalising constant to constrain $\sum_{j \geq 0} P(Z = j) = 1$.

We define Y as the number of secondary cases that both them and all their descendants are unsampled. Specifically,

$P(Y = k) = (\sum_{c \geq 1} P(C = c)(1 - \pi)^c)^k$ for $k$>0

$$P(Y = 0) = 1 - P(Y \geq 1)$$

where $\pi$ is the reporting probability and $P(C)$ is the probability distribution of the number of cases in a cluster of descendants (including itself). Specifically, $P(C)$ can be defined in terms of the number of true secondary cases, Z, as:

$$P(C = 1) = P(Z = 0),$$

$$P(C = 2) = P(Z = 1),$$

$P(C = 3) = P(Z = 2) + P(Z = 1)^2$, etc.

Assuming $P(Z = k)(1 - \pi)^j \approx 0$when $j > 2, k > 1$, we can write,

$P(Y = i) = ((1 - \pi)P(Z = 0) + (1 - \pi)^2 P(Z = 1))^i$ for i>0

$$P(Y = 0) = 1 - P(Y \geq 1)$$

Now, we define

$$P(Y = k \mid p, q) = ((1 - \pi)p + (1 - \pi)^2 q)^k$$

$$P(Y = 0 \mid p, q) = 1 - P(Y \geq 1 \mid p, q)$$

and we generate samples from the two distributions P(X) and P(Y | $p$, $q$) to generate a sample of Z and, from this sample, we estimate P(Z | $p$, $q$). We then calculate the unique $p$ and $q$ that satisfies $P(Y = k \mid p, q) = ((1-\pi)P(Z=0) \quad + (1-\pi)^2 P(Z=1))^k$.

We therefore calculate a secondary case distribution, $P(Z_h = i)$ for each sampled transmission tree, $h$, in the posterior distribution.

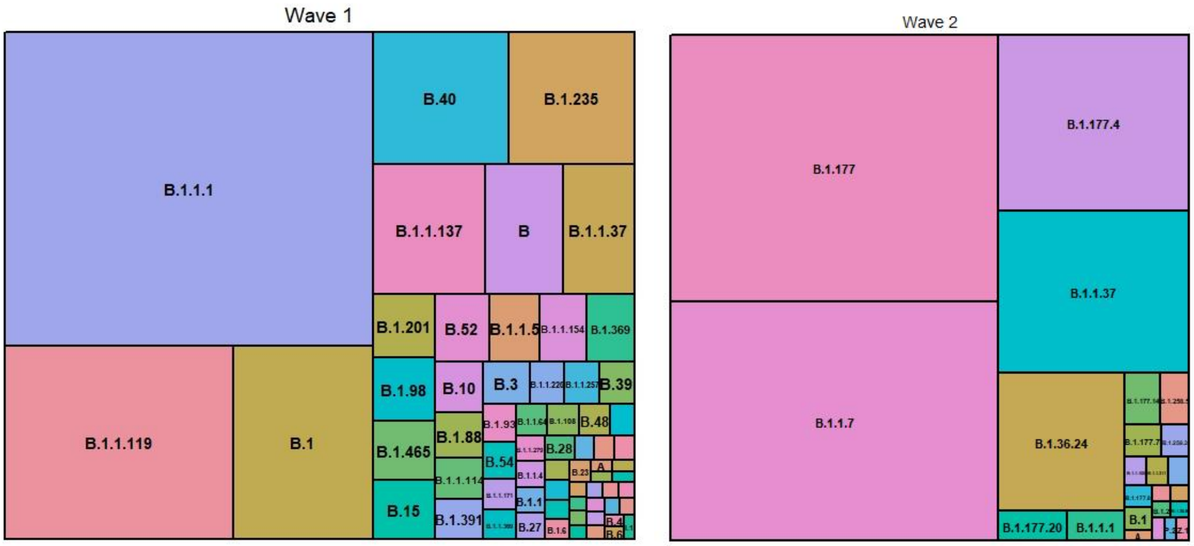


**Supplementary Figure S1. Treemap of all PANGO lineages represented in the Wave 1 and Wave 2 datasets.**

The size of each rectangle is proportional to the number of each lineage in the dataset.

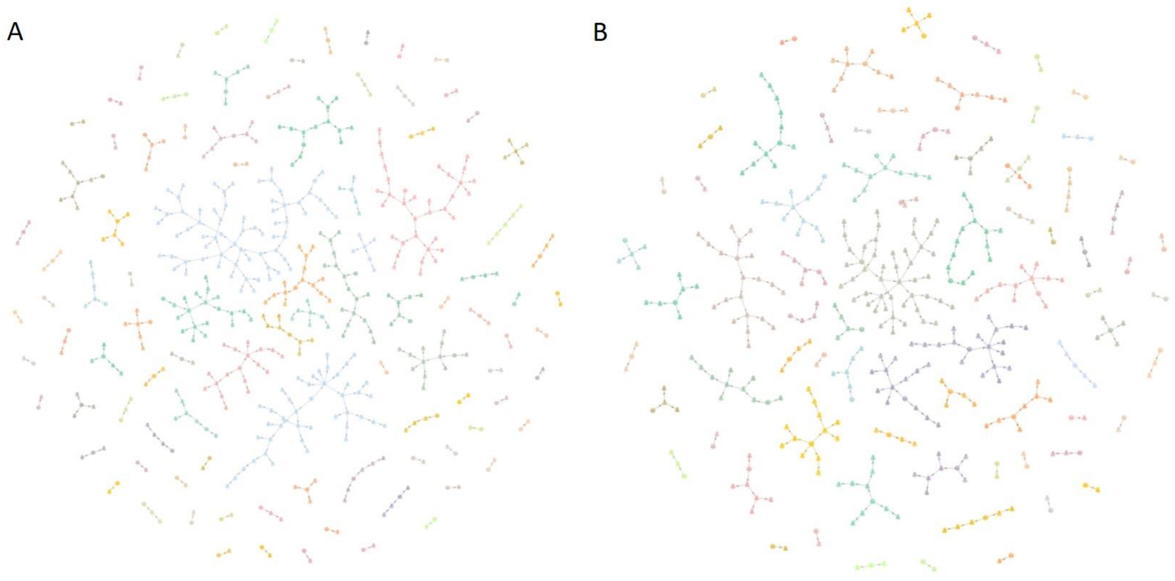


**Supplementary Figure S2. Network of all transmission chains in Wave 1 and 2**
(A) *All transmission chains from a representative network in* Wave 1. (B) *All transmission chains from a representative network in Wave 2. Each colour represents a different transmission chain.*

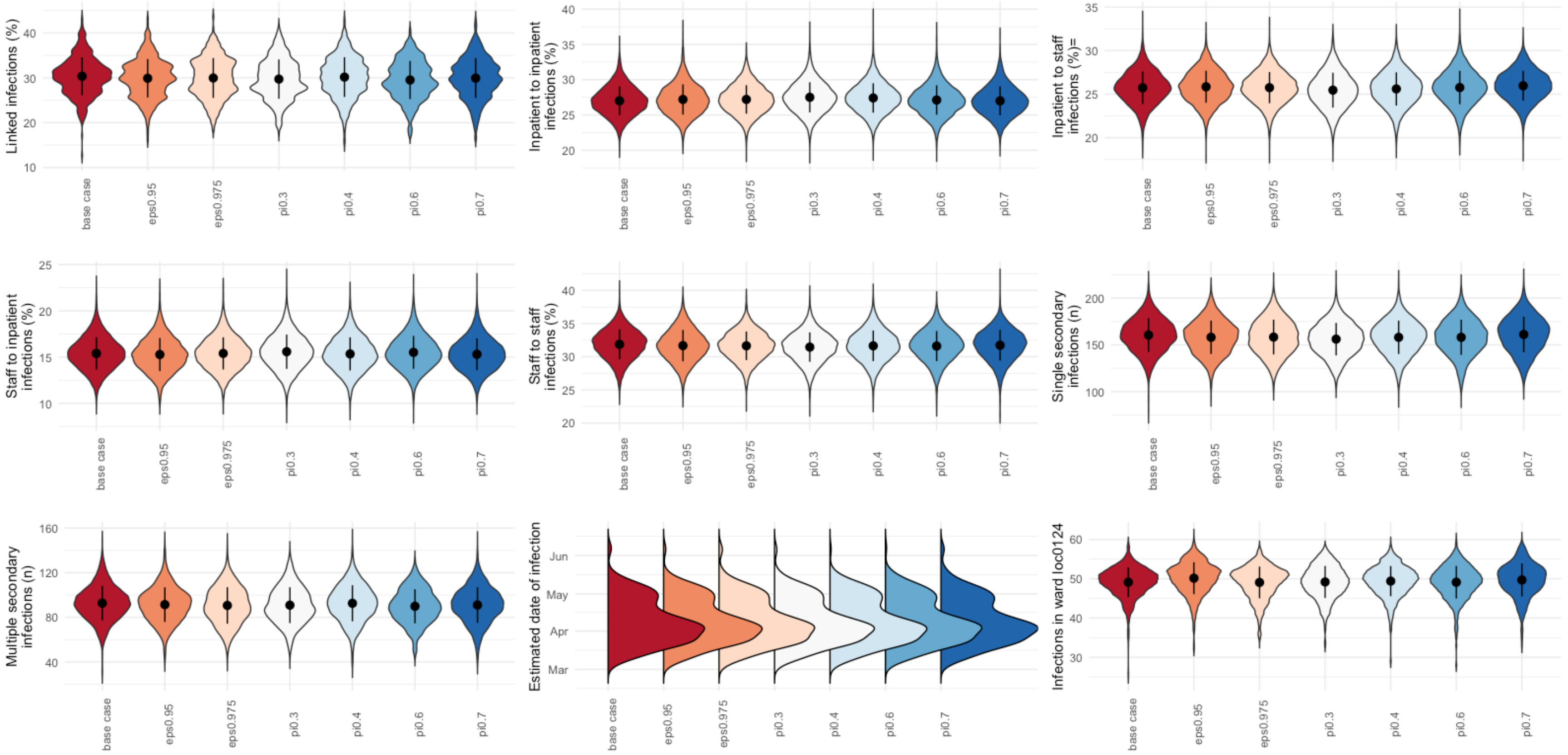

**Supplementary Figure S3. Sensitivity analysis of Wave 1.**

*eps ($\sigma$) and pi ($\pi$) refer to the probability of $\alpha_i$ and $i$ being registered on the same ward on the day of infection and the proportion of cases sampled in the outbreak, respectively (as in the notation Supplementary Table S1).*

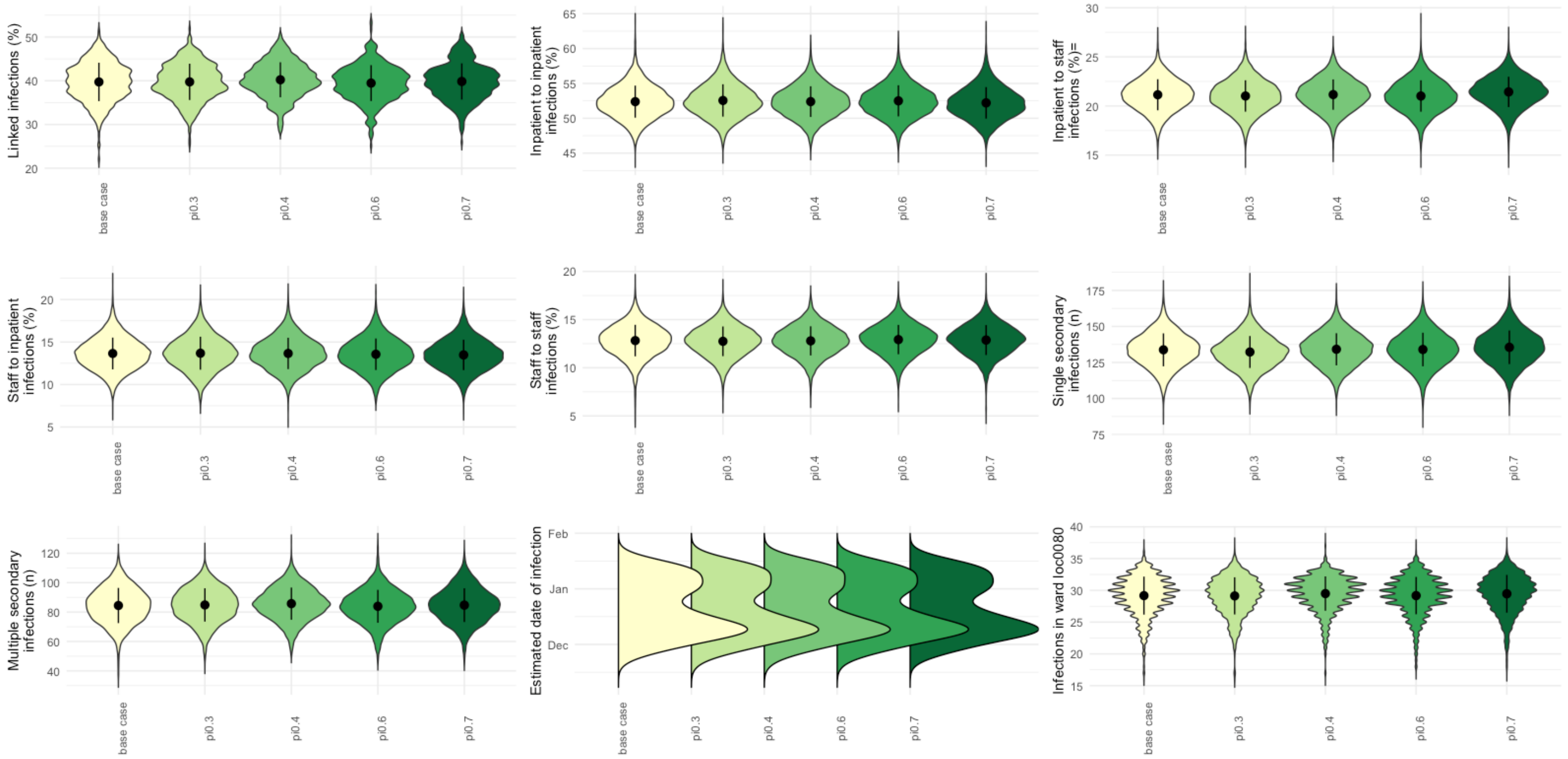


**Supplementary Figure S4. Sensitivity analysis of wave 2.**
*pi (π) refers to the proportion of cases sampled in the outbreak as in the notation Supplementary table S1.*

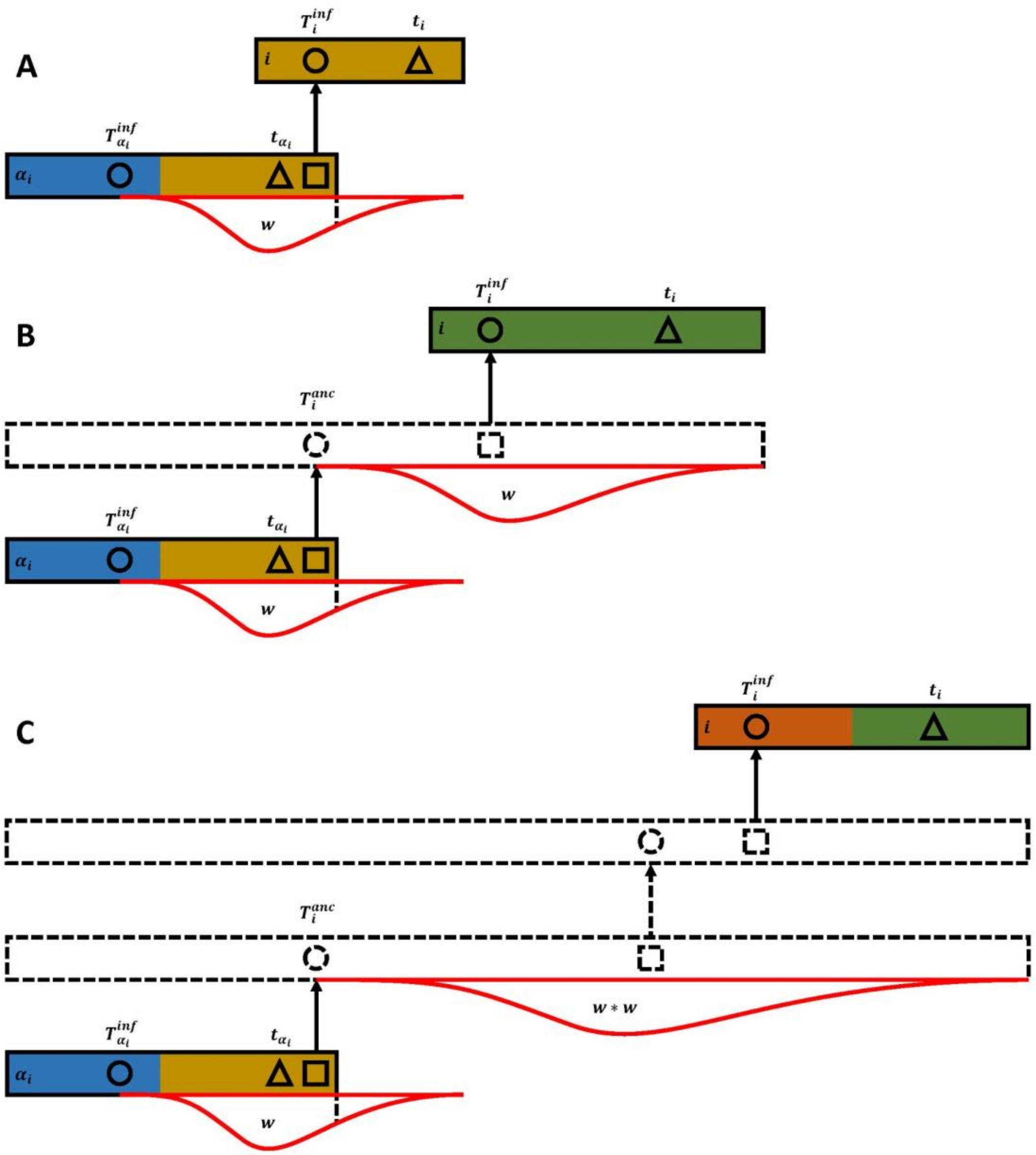


**Supplementary Figure S5. Schematic representation of the outbreaker ward model**

*Rows represent individual cases, colours indicate the ward a case is registered in, dotted outlines indicate unobserved cases. Circles represent infection times, triangles dates of symptom onset, squares dates of onward infection. Red curved lines indicate the generation time distribution. (A) A case i and its infector are both sampled. The wards of the infectee and infector are directly observed and the probability of infection between these wards on the day of infection can be directly calculated. (B) A case i and its most recent sampled ancestor $\alpha_i$ are separated by an unobserved case. The ancestral infection time $T_i^{anc}$ is imputed and the potential ward states of the unobserved case on the day of infection and day of onward infection are integrated over. (C) A case i and its most recent sampled ancestor $\alpha_i$ are separated by two unobserved cases. The ancestral infection time $T_i^{anc}$ is imputed and the ward states of both unobserved cases integrated over.*

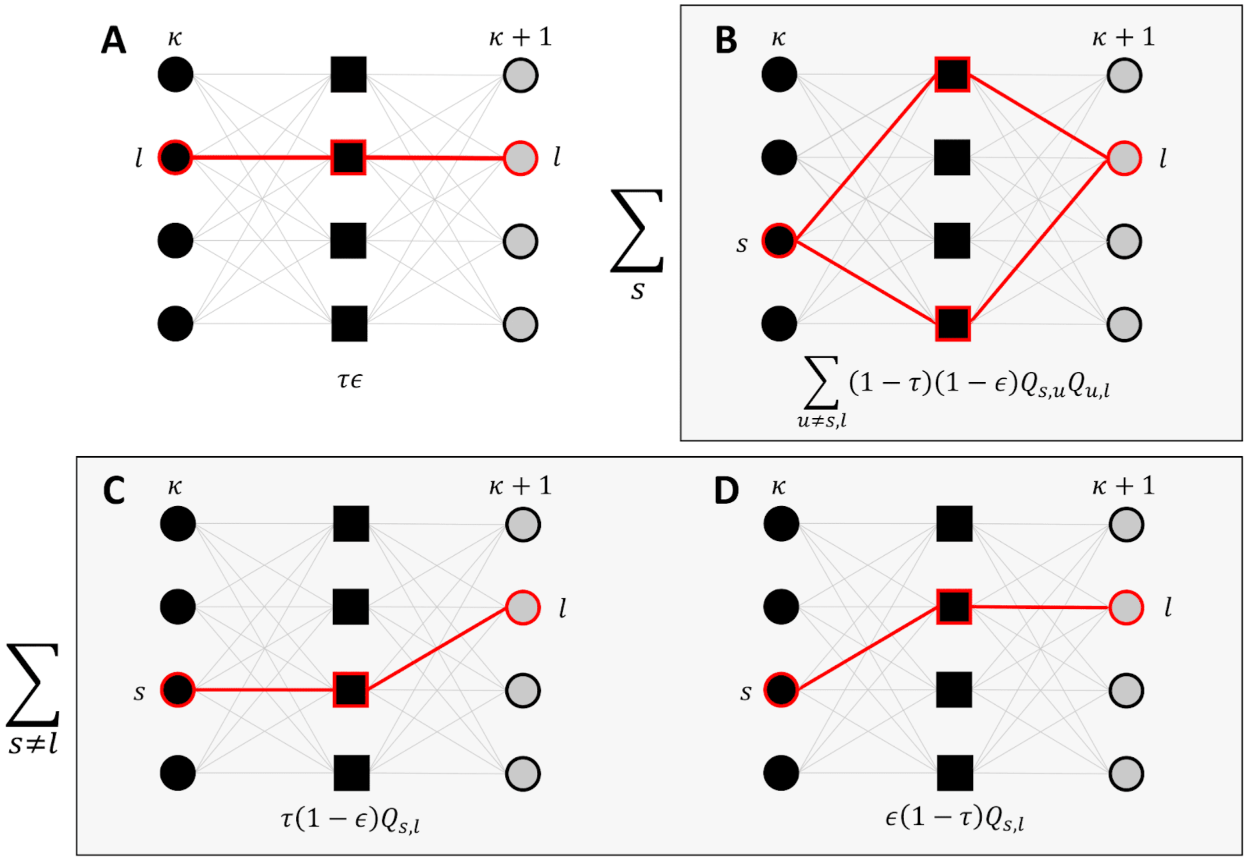


**Supplementary Figure S6. Integrating over transmission pathways to a given ward *l*.**
*Each row represents a ward. Black circles represent the ward the primary case is infected on. Black squares represent the ward of the primary case on the day of onward infection. Grey circles represent the ward the secondary case is infected on. A case remains on the ward it was infected on with probability $\tau$. An infection occurs within a ward with probability $\sigma$. All probabilities described in this figure are conditional on the primary case becoming infected on that given ward. A) The primary case is infected on ward l, remains on the same ward and infection occurs within the ward. B) The primary case is infected on any ward s, moves to an intermediate ward that is not s or l, and infects a case on ward l. The probability of this scenario must integrate over all potential initial wards s. C) The primary case is infected on a ward s that is not l, remains on ward s and causes infection in ward l. D) The primary case is infected on a ward s that is not l, moves to ward l and causes infection within ward l. The probability of scenarios C) and D) must integrate over all potential initial wards that are not l. Fast integration over the transmission pathways given in panels A-D can be achieved by the matrix multiplication* $Y(\tau) \times X(\sigma)$.

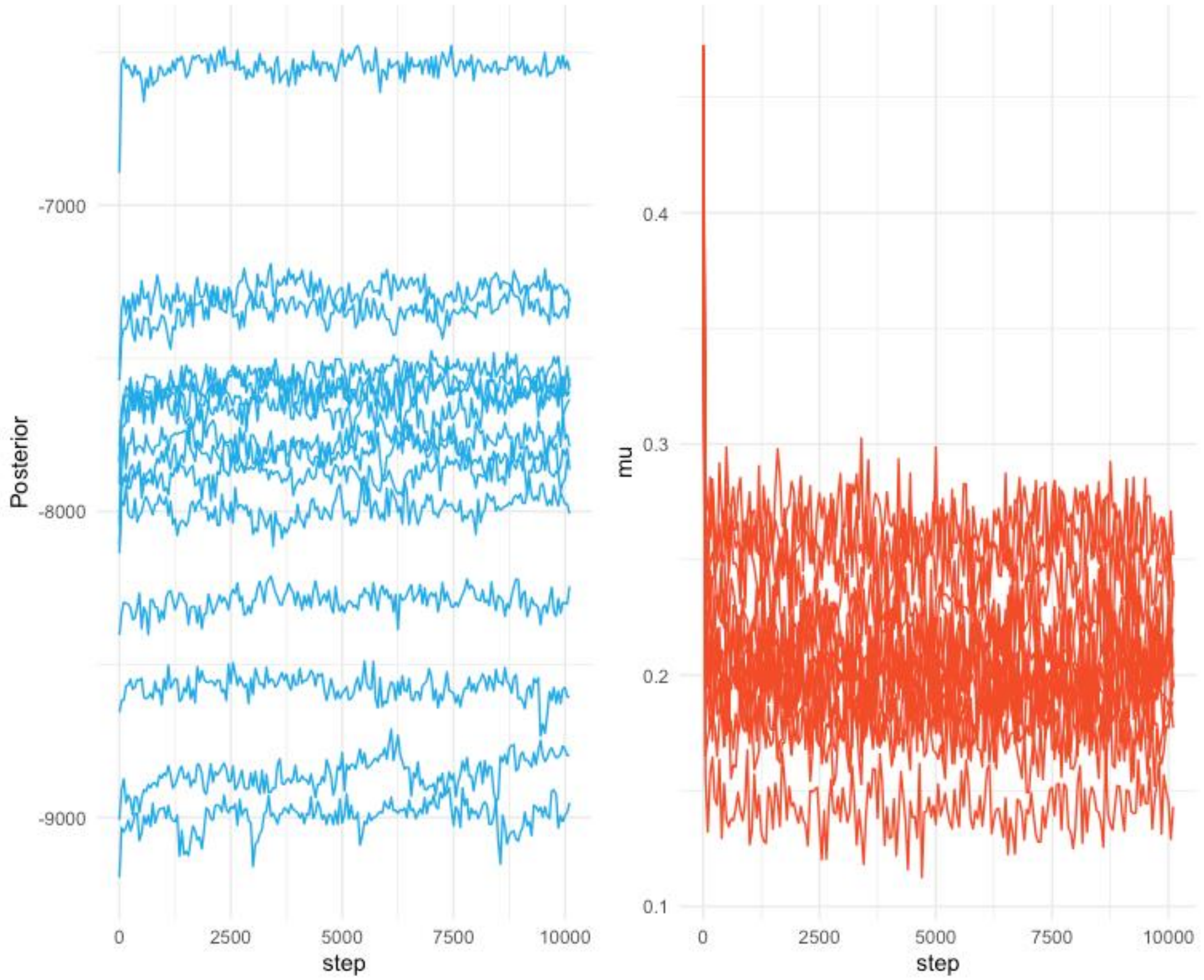


**Supplementary Figure S6. Trace of the log-posterior values and $\mu$ in exploratory runs**

### Supplementary Table S1. Notation of *outbreaker2* model

| Symbol | Type | Description | *Value (when applicable)* |
|---|---|---|---|
| $i$ | Data | Index of cases | |
| N | Data | Number of cases in the sample | |
| $s_i$ | Data | Sequence of case $i$ | |
| $t_i$ | Data | Collection date of $s_i$ | |
| $W_{i,t}$ | Data | Registered ward of case $i$ on day $t$ | |
| $w$ | Function | Generation time distribution | Discrete Gamma (shape =4.83, scale = 1.05)[5,6] |
| $f$ | Function | Incubation period distribution | Discrete Gamma (shape = 5.89, scale=1.07)[5,6] |
| $d(s_i, s_j)$ | Function | Number of mutations between $s_i$ and $s_j$ | |
| $l(s_i, s_j)$ | Function | Number of comparable nucleotide positions between $s_i$ and $s_j$ | |
| $\alpha_i$ | Augmented data | Index of the most recent sampled ancestor of case $i$ | |
| $\kappa_i$ | Augmented data | Number of generations between $\alpha_i$ and $i$ | |
| $T_i^{inf}$ | Augmented data | Date of infection of $i$ | |
| $T_i^{anc}$ | Augmented data | Date of infection of the earliest unobserved case in an unobserved transmission chain separating $\alpha_i$ and $i$ | |
| $\mu$ | Parameter | Mutation rate of SARS-CoV-2, per site per generation of infection | Beta ($\alpha$ = 89.9, $\beta$ = 809.1)[7] |
| $\pi$ | Parameter | Proportion of cases sampled in the outbreak | 0.5 |
| $\sigma$ | Parameter | Probability of $\alpha_i$ and i being registered on the same ward on the day of infection | 0.99 |
| $\tau$ | Parameter | Daily probability of an unobserved case being registered on the same ward it was in on the day of infection | |

**Supplementary Table S2. Percentage of each onset and association with infectors or onward secondary infections in the outbreaker model**

| | Assigned infector (hospital acquired infections) % (95% Credible interval) | | Secondary infections % (95% Credible interval) | |
|---|---|---|---|---|
| | **Wave 1** | **Wave 2** | **Wave 1** | **Wave 2** |
| Community onset community associated | 0 | 0 | 3.9 (2.1-6.0) | 3.5 (1.7-5.9) |
| Community onset suspected healthcare associated | 35.0 (24.1-45.6) | 42.2 (31.3-52.2) | 18.1 (10.1-26.6) | 24.0 (14.9-32.8) |
| Hospital onset indeterminate hospital acquired | 32.1 (21.3-41.0) | 52.3 (41.6-61.8) | 27.0 (18.9-35.1) | 39.1 (29.5-47.7) |
| Hospital onset suspected hospital acquired | 71.3 (56.8-82.4) | 83.6 (69.3-93.2) | 59.6 (45.9-72.1) | 52.4 (42.7-61.8) |
| Hospital onset hospital acquired | 69.8 (55.0-80.0) | 77.4 (61.0-89.0) | 45.2 (35.0-55.0) | 51.2 (41.0-60.0) |

**Supplementary File S3: Supplementary Authors**

Sheffield COVID-19 Genomics Group

Alison Cope, Nasar Ali, Rasha Raghei, Joe Heffer, Nikki Smith, Max Whiteley, Manoj Pohare, Samantha E. Hansford, Luke R. Green, Dennis Wang, Michael Anckorn, Adrienn Angyal, Rebecca Brown, Hailey Hornsby, Mehmet Yavuz, Danielle C. Groves, Paul J. Parsons, Rachel M. Tucker, Magdalena B. Dabrowska, Thomas Saville, Jose Schutter and Matthew D. Wyles

CMMID COVID-19 Working Group

Mihaly Koltai, Kerry LM Wong, Joel Hellewell, Emilie Finch, Matthew Quaife, James D Munday, Timothy W Russell, Kaja Abbas, Oliver Brady, Nicholas G. Davies, Alicia Rosello, Gwenan M Knight, Simon R Procter, William Waites, Nikos I Bosse, Stefan Flasche, Paul Mee, Rachel Lowe, Kathleen O'Reilly, Carl AB Pearson, Christopher I Jarvis, W John Edmunds, Fiona Yueqian Sun, Amy Gimma, Samuel Clifford, Rosanna C Barnard, Rachael Pung, Kiesha Prem, Sam Abbott, Sebastian Funk, Akira Endo, Sophie R Meakin, Yang Liu, Adam J Kucharski, Graham Medley, Ciara V McCarthy, Yalda Jafari, Mark Jit, Damien C Tully, Hamish P Gibbs, Lloyd A C Chapman, Billy J Quilty, Frank G Sandmann, David Hodgson and Rosalind M Eggo.

The COVID-19 Genomics UK (COG-UK) consortium

**Funding acquisition, Leadership and supervision, Metadata curation, Project administration, Samples and logistics, Sequencing and analysis, Software and analysis tools, and Visualisation:**

Dr Samuel C Robson PhD [13].

**Funding acquisition, Leadership and supervision, Metadata curation, Project administration, Samples and logistics, Sequencing and analysis, and Software and analysis tools:**

Prof Nicholas J Loman PhD [41] and Dr Thomas R Connor PhD [10, 69].

**Leadership and supervision, Metadata curation, Project administration, Samples and logistics, Sequencing and analysis, Software and analysis tools, and Visualisation:**

Dr Tanya Golubchik PhD [5].

**Funding acquisition, Metadata curation, Samples and logistics, Sequencing and analysis, Software and analysis tools, and Visualisation:**

Dr Rocio T Martinez Nunez PhD [42].

**Funding acquisition, Leadership and supervision, Metadata curation, Project administration, and Samples and logistics:**

Dr Catherine Ludden PhD [88].

**Funding acquisition, Leadership and supervision, Metadata curation, Samples and logistics, and Sequencing and analysis:**

Dr Sally Corden PhD [69].

**Funding acquisition, Leadership and supervision, Project administration, Samples and logistics, and Sequencing and analysis:**

Ian Johnston [99] and Dr David Bonsall PhD [5].

**Funding acquisition, Leadership and supervision, Sequencing and analysis, Software and analysis tools, and Visualisation:**

Prof Colin P Smith PhD [87] and Dr Ali R Awan PhD [28].

**Funding acquisition, Samples and logistics, Sequencing and analysis, Software and analysis tools, and Visualisation:**

Dr Giselda Bucca PhD [87].

**Leadership and supervision, Metadata curation, Project administration, Samples and logistics, and Sequencing and analysis:**

Dr M. Estee Torok FRCP [22, 101].

**Leadership and supervision, Metadata curation, Project administration, Samples and logistics, and Visualisation:**

Dr Kordo Saeed MD/ FRCPath [81, 110] and Dr Jacqui A Prieto PhD [83, 109].

**Leadership and supervision, Metadata curation, Project administration, Sequencing and analysis, and Software and analysis tools:**

Dr David K Jackson PhD [99].

**Metadata curation, Project administration, Samples and logistics, Sequencing and analysis, and Software and analysis tools:**

Dr William L Hamilton PhD [22].

**Metadata curation, Project administration, Samples and logistics, Sequencing and analysis, and Visualisation:**

Dr Luke B Snell MSc/ MBBS [11].

**Funding acquisition, Leadership and supervision, Metadata curation, and Samples and logistics:**

Dr Catherine Moore [69].

**Funding acquisition, Leadership and supervision, Project administration, and Samples and logistics:**

Dr Ewan M Harrison PhD [99, 88].

**Leadership and supervision, Metadata curation, Project administration, and Samples and logistics:**

Dr Sonia Goncalves PhD [99] and Dr Leigh M Jackson Ph.D. [91].

**Leadership and supervision, Metadata curation, Samples and logistics, and Sequencing and analysis:**

Prof Ian G Goodfellow PhD [24], Dr Derek J Fairley PhD [3, 72], Prof Matthew W Loose PhD [18] and Joanne Watkins MSc [69].

**Leadership and supervision, Metadata curation, Samples and logistics, and Software and analysis tools:**

Rich Livett MSc [99].

**Leadership and supervision, Metadata curation, Samples and logistics, and Visualisation:**

Dr Samuel Moses MD [25, 106].

**Leadership and supervision, Metadata curation, Sequencing and analysis, and Software and analysis tools:**

Dr Roberto Amato PhD [99], Dr Sam Nicholls PhD [41] and Dr Matthew Bull PhD [69].

**Leadership and supervision, Project administration, Samples and logistics, and Sequencing and analysis:**

Prof Darren L Smith PhD [37, 58, 105].

**Leadership and supervision, Sequencing and analysis, Software and analysis tools, and Visualisation:**

Dr Jeff Barrett PhD [99] and Prof David M Aanensen PhD [14, 114].

**Metadata curation, Project administration, Samples and logistics, and Sequencing and analysis:**

Dr Martin D Curran PhD [65], Dr Surendra Parmar PhD [65], Dr Dinesh Aggarwal MRCP [95, 99, 64] and Dr James G Shepherd MBChB/MRCP [48].

**Metadata curation, Project administration, Sequencing and analysis, and Software and analysis tools:**

Dr Matthew D Parker PhD [93].

**Metadata curation, Samples and logistics, Sequencing and analysis, and Visualisation:**

Dr Sharon Glaysher PhD [61].

**Metadata curation, Sequencing and analysis, Software and analysis tools, and Visualisation:**

Dr Matthew Bashton PhD [37, 58], Dr Anthony P Underwood PhD [14, 114], Dr Nicole Pacchiarini PhD [69] and Dr Katie F Loveson PhD [77].

**Project administration, Sequencing and analysis, Software and analysis tools, and Visualisation:**

Dr Alessandro M Carabelli PhD [88].

**Funding acquisition, Leadership and supervision, and Metadata curation:**

Dr Kate E Templeton PhD [53, 90].

**Funding acquisition, Leadership and supervision, and Project administration:**

Dr Cordelia F Langford PhD [99], John Sillitoe BEng [99], Dr Thushan I de Silva PhD [93] and Dr Dennis Wang PhD [93].

**Funding acquisition, Leadership and supervision, and Sequencing and analysis:**

Prof Dominic Kwiatkowski [99, 107], Prof Andrew Rambaut DPhil [90], Dr Justin O'Grady PhD [70, 89] and Dr Simon Cottrell PhD [69].

**Leadership and supervision, Metadata curation, and Sequencing and analysis:**

Prof Matthew T.G. Holden PhD [68] and Prof Emma C Thomson PhD/FRCP [48].

**Leadership and supervision, Project administration, and Samples and logistics:**

Dr Husam Osman PhD [64, 36], Dr Monique Andersson PhD [59], Prof Anoop J Chauhan [61] and Dr Mohammed O Hassan-Ibrahim PhD/FRCPath [6].

**Leadership and supervision, Project administration, and Sequencing and analysis:**

Dr Mara Lawniczak [99].

**Leadership and supervision, Samples and logistics, and Sequencing and analysis:**

Prof Ravi Kumar Gupta PhD [88, 113], Dr Alex Alderton PhD [99], Dr Meera Chand [66], Dr Chrystala Constantinidou PhD [94], Dr Meera Unnikrishnan PhD [94], Prof Alistair C Darby PhD [92], Prof Julian A Hiscox PhD [92] and Prof Steve Paterson PhD [92].

**Leadership and supervision, Sequencing and analysis, and Software and analysis tools:**

Dr Inigo Martincorena [99], Prof David L Robertson PhD [48], Dr Erik M Volz PhD [39], Dr Andrew J Page PhD [7] and Prof Oliver G Pybus DPhil [23].

**Leadership and supervision, Sequencing and analysis, and Visualisation:**

Dr Andrew R Bassett PhD [99].

**Metadata curation, Project administration, and Samples and logistics:**

Dr Cristina V Ariani PhD [99], Dr Michael H Spencer Chapman MBBS [99, 88], Dr Kathy K Li MBBCh/FRCPath [48], Dr Rajiv N Shah BMBS/MRCP/MSc [48], Dr Natasha G Jesudason MBChB MRCP FRCPath [48] and Dr Yusri Taha MD/PhD [50].

**Metadata curation, Project administration, and Sequencing and analysis:**

Martin P McHugh MSc [53], Dr Rebecca Dewar PhD [53].

**Metadata curation, Samples and logistics, and Sequencing and analysis:**

Dr Aminu S Jahun PhD [24], Dr Claire McMurray PhD [41], Ms Sarojini Pandey MSc [84], Dr James P McKenna PhD [3], Dr Andrew Nelson PhD [58, 105], Dr Gregory R Young PhD [37, 58], Dr Clare M McCann PhD [58, 105] and Mr Scott Elliott [61].

**Metadata curation, Samples and logistics, and Visualisation:**

Ms Hannah Lowe MSc [25].

**Metadata curation, Sequencing and analysis, and Software and analysis tools:**

Dr Ben Temperton Ph.D. [91], Dr Sunando Roy PhD [82], Dr Anna Price PhD [10], Dr Sara Rey PhD [69] and Mr Matthew Wyles [93].

**Metadata curation, Sequencing and analysis, and Visualisation:**

Stefan Rooke MSc [90] and Dr Sharif Shaaban PhD [68].

**Project administration, Samples and logistics, Sequencing and analysis:**

Dr Mariateresa de Cesare PhD [98].

**Project administration, Samples and logistics, and Software and analysis tools:**

Laura Letchford BSc [99].

**Project administration, Samples and logistics, and Visualisation:**

Miss Siona Silveira MSc [81], Dr Emanuela Pelosi FRCPath [81] and Dr Eleri Wilson-Davies MD/FRCPath [81].

**Samples and logistics, Sequencing and analysis, and Software and analysis tools:**

Dr Myra Hosmillo PhD [24].

**Sequencing and analysis, Software and analysis tools, and Visualisation:**

Áine O'Toole MSc [90], Dr Andrew R Hesketh PhD [87], Mr Richard Stark MSc [94], Dr Louis du Plessis PhD [23], Dr Chris Ruis PhD [88], Dr Helen Adams PhD [4] and Dr Yann Bourgeois PhD [76].

**Funding acquisition, and Leadership and supervision:**

Dr Stephen L Michell PhD [91], Prof Dimitris Grammatopoulos PhD/FRCPath [84, 112], Dr Jonathan Edgeworth PhD/FRCPath [12], Prof Judith Breuer MD [30, 82], Prof John A Todd PhD [98] and Dr Christophe Fraser PhD [5].

**Funding acquisition, and Project administration:**

Dr David Buck PhD [98] and Michaela John BSc [9].

**Leadership and supervision, and Metadata curation:**

Dr Gemma L Kay PhD [70].

**Leadership and supervision, and Project administration:**

Steve Palmer [99], Prof Sharon J Peacock [88, 64] and David Heyburn [69].

**Leadership and supervision, and Samples and logistics:**

Danni Weldon BSc [99], Dr Esther Robinson PhD [64, 36], Prof Alan McNally PhD [41, 86], Dr Peter Muir PhD [64], Dr Ian B Vipond PhD [64], Dr John BoYes MBChB [29], Dr Venkat Sivaprakasam PhD [46], Dr Tranprit Saluja FRCPath/MD [75], Dr Samir Dervisevic FRCPath [54] and Dr Emma J Meader FRCPath [54].

**Leadership and supervision, and Sequencing and analysis:**

Dr Naomi R Park PhD [99], Karen Oliver BSc [99], Dr Aaron R Jeffries Ph.D. [91], Dr Sascha Ott PhD [94], Dr Ana da Silva Filipe PhD [48], Dr David A Simpson PhD [72] and Dr Chris Williams MB BS [69].

**Leadership and supervision, and Visualisation:**

Dr Jane AH Masoli MBChB [73, 91].

**Metadata curation, and Samples and logistics:**

Dr Bridget A Knight PhD. [73, 91], Dr Christopher R Jones Ph.D. [73, 91], Mr Cherian Koshy MSc CSci FIBMS [1], Miss Amy Ash BSc [1], Dr Anna Casey PhD [71], Dr Andrew Bosworth PhD [64, 36], Dr Liz Ratcliffe PhD [71], Dr Li Xu-McCrae PhD [36], Miss Hannah M Pymont MSc [64], Ms Stephanie Hutchings [64], Dr Lisa Berry PhD [84], Ms Katie Jones MSc [84], Dr Fenella Halstead PhD [46], Mr Thomas Davis MSc [21], Dr Christopher Holmes PhD [16], Prof Miren Iturriza-Gomara PhD [92], Dr Anita O Lucaci PhD [92], Dr Paul Anthony Randell MBBCh [38, 104], Dr Alison Cox PhD [38, 104], Pinglawathee Madona [38, 104], Dr Kathryn Ann Harris PhD [30], Dr Julianne Rose Brown PhD [30], Dr Tabitha W Mahungu FRCPath [74], Dr Dianne Irish-Tavares FRCPath [74], Dr Tanzina Haque

FRCPath PhD [74], Dr Jennifer Hart MRCP [74], Mr Eric Witele MSc [74], Mrs Melisa Louise Fenton DipHE [75], Mr Steven Liggett [79], Dr Clive Graham MD [56], Ms Emma Swindells Bsc [57], Ms Jennifer Collins BSc [50], Mr Gary Eltringham BSc [50], Ms Sharon Campbell MSc [17], Dr Patrick C McClure PhD [97], Dr Gemma Clark PhD [15], Dr Tim J Sloan PhD [60], Mr Carl Jones [15] and Dr Jessica Lynch PhD MBChB [2, 111].

**Metadata curation, and Sequencing and analysis:**

Dr Ben Warne MRCP [8], Steven Leonard PhD [99], Jillian Durham BSc [99], Dr Thomas Williams MD [90], Dr Sam T Haldenby PhD [92], Dr Nathaniel Storey PhD [30], Dr Nabil-Fareed Alikhan PhD [70], Dr Nadine Holmes PhD [18], Dr Christopher Moore PhD [18], Mr Matthew Carlile BSc [18], Malorie Perry MSc [69], Dr Noel Craine DPhil [69], Prof Ronan A Lyons MD [80], Miss Angela H Beckett MSc [13], Salman Goudarzi PhD [77], Christopher Fearn MRes [77], Kate Cook [77], Hannah Dent BSc [77] and Hannah Paul MRes [77].

**Metadata curation, and Software and analysis tools:**

Robert Davies [99].

**Project administration, and Samples and logistics:**

Beth Blane BSc [88], Sophia T Girgis MSc [88], Dr Mathew A Beale PhD [99], Katherine L Bellis [99, 88], Matthew J Dorman [99], Eleanor Drury [99], Leanne Kane [99], Sally Kay [99], Dr Samantha McGuigan [99], Dr Rachel Nelson PhD [99], Liam Prestwood [99], Dr Shavanthi Rajatileka PhD [99], Dr Rahul Batra MD [12], Dr Rachel J Williams PhD [82], Dr Mark Kristiansen PhD [82], Dr Angie Green PhD [98], Miss

Anita Justice MSc [59], Dr Adhyana I.K Mahanama MD [81, 102] and Dr Buddhini Samaraweera MD [81, 102].

**Project administration, and Sequencing and analysis:**

Dr Nazreen F Hadjirin PhD [88] and Dr Joshua Quick PhD [41].

**Project administration, and Software and analysis tools:**

Mr Radoslaw Poplawski BSc [41].

**Samples and logistics, and Sequencing and analysis:**

Leanne M Kermack MSc [88], Nicola Reynolds PhD [7], Grant Hall BS [24], Yasmin Chaudhry BSc [24], Malte L Pinckert MPhil [24], Dr Iliana Georgana PhD [24], Dr Robin J Moll PhD [99], Dr Alicia Thornton [66], Dr Richard Myers [66], Dr Joanne Stockton PhD [41], Miss Charlotte A Williams BSc [82], Dr Wen C Yew PhD [58], Alexander J Trotter MRes [70], Miss Amy Trebes MSc [98], Mr George MacIntyre-Cockett BSc [98], Alec Birchley MSc [69], Alexander Adams BSc [69], Amy Plimmer [69], Bree Gatica-Wilcox MPhil [69], Dr Caoimhe McKerr PhD [69], Ember Hilvers MA [69], Hannah Jones [69], Dr Hibo Asad PhD [69], Jason Coombes BSc [69], Johnathan M Evans MSc [69], Laia Fina [69], Lauren Gilbert A-Levels [69], Lee Graham BSc [69], Michelle Cronin [69], Sara Kumziene-SummerhaYes MSc [69], Sarah Taylor [69], Sophie Jones MSc [69], Miss Danielle C Groves BA [93], Mrs Peijun Zhang MSc [93], Miss Marta Gallis MSc [93] and Miss Stavroula F Louka MSc [93].

**Samples and logistics, and Software and analysis tools:**

Dr Igor Starinskij Msc MRCP [48].

**Sequencing and analysis, and Software and analysis tools:**

Dr Chris J Illingworth PhD [47], Dr Chris Jackson PhD [47], Ms Marina Gourtovaia MSc [99], Gerry Tonkin-Hill [99], Kevin Lewis [99], Dr Jaime M Tovar-Corona PhD [99], Dr Keith James PhD [99], Dr Laura Baxter PhD [94], Dr Mohammad T. Alam PhD [94], Dr Richard J Orton PhD [48], Dr Joseph Hughes PhD [48], Dr Sreenu Vattipally PhD [48], Dr Manon Ragonnet-Cronin PhD [39], Dr Fabricia F. Nascimento PhD [39], Mr David Jorgensen MSc [39], Ms Olivia Boyd MSc [39], Ms Lily Geidelberg MSc [39], Dr Alex E Zarebski PhD [23], Dr Jayna Raghwani PhD [23], Dr Moritz UG Kraemer DPhil [23], Joel Southgate MSc [10, 69], Dr Benjamin B Lindsey MRCP [93] and Mr Timothy M Freeman MPhil [93].

**Software and analysis tools, and Visualisation**:

Jon-Paul Keatley [99], Dr Joshua B Singer PhD [48], Leonardo de Oliveira Martins PhD [70], Dr Corin A Yeats PhD [14], Dr Khalil Abudahab PhD [14, 114], Mr Ben EW Taylor MEng [14, 114] and Mirko Menegazzo [14].

**Leadership and supervision**:

Prof John Danesh [99], Wendy Hogsden MSc [46], Dr Sahar Eldirdiri MBBS MSc FRCPath [21], Mrs Anita Kenyon MSc [21], Dr Jenifer Mason MBBS [43], Mr Trevor I Robinson MSc [43], Prof Alison Holmes MD [38, 103], Dr James Price PhD [38, 103], Prof John A Hartley PhD [82], Dr Tanya Curran PhD [3], Dr Alison E Mather PhD [70], Dr Giri Shankar [69], Dr Rachel Jones [69], Dr Robin Howe [69] and Dr Sian Morgan FRCPath [9].

**Metadata curation:**

Dr Elizabeth Wastenge MD [53], Dr Michael R Chapman PhD [34, 88, 99], Mr Siddharth Mookerjee MPH [38, 103], Dr Rachael Stanley PhD [54], Mrs Wendy Smith [15], Prof Timothy Peto PhD [59], Dr David Eyre PhD [59], Dr Derrick Crook [59], Dr Gabrielle Vernet MBBS [33], Dr Christine Kitchen PhD [10], Huw Gulliver [10], Dr Ian Merrick PhD [10], Prof Martyn Guest PhD [10], Robert Munn BSc [10], Dr Declan T Bradley [63, 72] and Dr Tim Wyatt [63].

**Project administration:**

Dr Charlotte Beaver [99], Luke Foulser [99], Sophie Palmer [88], Carol M Churcher [88], Ellena Brooks MA [88], Kim S Smith [88], Dr Katerina Galai PhD [88], Georgina M McManus BSc [88], Dr Frances Bolt PhD [38, 103], Dr Francesc Coll PhD [19], Lizzie Meadows MA [70], Dr Stephen W Attwood PhD [23], Dr Alisha Davies [69], Elen De Lacy MSc [69], Fatima Downing [69], Sue Edwards [69], Dr Garry P Scarlett PhD [76], Mrs Sarah Jeremiah MSc [83] and Dr Nikki Smith PhD [93].

**Samples and logistics:**

Danielle Leek Bsc [88], Sushmita Sridhar BS [88, 99], Sally Forrest BSc [88], Claire Cormie [88], Harmeet K Gill PhD [88], Joana Dias MSc [88], Ellen E Higginson PhD [88], Mailis Maes MPhil [88], Jamie Young BSc [88], Michelle Wantoch PhD [7], Sanger Covid Team (www.sanger.ac.uk/covid-team) [99], Dorota Jamrozy [99], Stephanie Lo [99], Dr Minal Patel PhD [99], Verity Hill [90], Ms Claire M Bewshea MSc [91], Prof Sian Ellard FRCPath [73, 91], Dr Cressida Auckland FRCPath [73], Dr Ian Harrison [66], Dr Chloe Bishop [66], Dr Vicki Chalker [66], Dr Alex Richter PhD [85], Dr Andrew Beggs PhD [85], Dr Angus Best PhD [86], Dr Benita Percival PhD [86], Dr Jeremy Mirza PhD [86], Dr Oliver Megram PhD [86], Dr Megan Mayhew PhD [86], Dr Liam Crawford PhD [86], Dr Fiona Ashcroft PhD [86], Dr Emma Moles-

Garcia PhD [86], Dr Nicola Cumley PhD [86], Mr Richard Hopes [64], Dr Patawee Asamaphan PhD [48], Mr Marc O Niebel MSc [48], Prof Rory N Gunson PhD FRCPath [100], Dr Amanda Bradley PhD [52], Dr Alasdair Maclean PhD [52], Dr Guy Mollett MBChB [52], Dr Rachel Blacow MBChB [52], Mr Paul Bird MSc [16], Mr Thomas Helmer [16], Miss Karlie Fallon [16], Dr Julian Tang [16], Dr Antony D Hale MBBS [49], Dr Louissa R Macfarlane-Smith PhD [49], Katherine L Harper MBiol [49], Miss Holli Carden MSc [49], Dr Nicholas W Machin MSc [45, 64], Ms Kathryn A Jackson MSc [92], Dr Shazaad S Y Ahmad MSc [45, 64], Dr Ryan P George PhD [45], Dr Lance Turtle PhD MRCP [92], Mrs Elaine O'Toole BSc [43], Mrs Joanne Watts BSc [43], Mrs Cassie Breen BSc [43], Mrs Angela Cowell MSc [43], Ms Adela Alcolea-Medina [32, 96], Ms Themoula Charalampous MSc [12, 42], Amita Patel [11], Dr Lisa J Levett PhD [35], Dr Judith Heaney PhD [35], Dr Aileen Rowan PhD [39], Prof Graham P Taylor DSc [39], Dr Divya Shah PhD [30], Miss Laura Atkinson MSc [30], Mr Jack CD Lee MSc [30], Mr Adam P Westhorpe BSc [82], Dr Riaz Jannoo PhD [82], Dr Helen L Lowe PhD [82], Miss Angeliki Karamani MSc [82], Miss Leah Ensell BSc [82], Mrs Wendy Chatterton MSc [35], Miss Monika Pusok MSc [35], Mrs Ashok Dadrah MSc [75], Miss Amanda Symmonds MSc [75], Dr Graciela Sluga MD/MSC [44], Dr Zoltan Molnar PhD [72], Mr Paul Baker MD [79], Prof Stephen Bonner [79], Ms Sarah Essex [79], Dr Edward Barton MD [56], Ms Debra Padgett BSc [56], Ms Garren Scott BSc [56], Ms Jane Greenaway MSc [57], Dr Brendan AI Payne MD [50], Dr Shirelle Burton-Fanning MD [50], Dr Sheila Waugh MD [50], Dr Veena Raviprakash MD [17], Ms Nicola Sheriff BSc [17], Ms Victoria Blakey BSc [17], ms Lesley-Anne Williams BSc [17], Dr Jonathan Moore MD [27], Ms Susanne Stonehouse BSc [27], Dr Louise Smith [55], Dr Rose K Davidson PhD [89], Dr Luke Bedford [26], Dr Lindsay Coupland PhD [54], Ms Victoria Wright BSc [18], Dr Joseph G Chappell PhD [97], Dr Theocharis Tsoleridis PhD [97], Prof Jonathan Ball PhD [97], Mrs Manjinder Khakh [15], Dr Vicki M Fleming PhD [15], Dr Michelle M Lister PhD [15], Dr Hannah C Howson-Wells PhD [15], Dr Louise Berry [15], Dr Tim Boswell [15], Dr Amelia Joseph [15],

Dr Iona Willingham [15], Dr Nichola Duckworth [60], Dr Sarah Walsh [60], Dr Emma Wise PhD [2, 111], Dr Nathan Moore PhD [2, 111], Miss Matilde Mori BSc [2, 108, 111], Dr Nick Cortes MRCP FRCPath [2, 111], Dr Stephen Kidd PhD [2, 111], Dr Rebecca Williams BMBS [33], Laura Gifford MSc [69], Miss Kelly Bicknell [61], Dr Sarah Wyllie [61], Miss Allyson Lloyd [61], Mr Robert Impey MSc [61], Ms Cassandra S Malone MSc [6], Mr Benjamin J Cogger BSc [6], Nick Levene MSc [62], Lynn Monaghan [62], Dr Alexander J Keeley MRCP [93], Dr David G Partridge FRCP FRCPath [78, 93], Dr Mohammad Raza [78, 93], Dr Cariad Evans [78, 93] and Dr Kate Johnson [78, 93].

**Sequencing and analysis:**

Emma Betteridge BSc [99], Ben W Farr BSc [99], Scott Goodwin MSc [99], Dr Michael A Quail PhD [99], Carol Scott [99], Lesley Shirley MSc [99], Scott AJ Thurston BSc [99], Diana Rajan MSc [99], Dr Iraad F Bronner PhD [99], Louise Aigrain PhD [99], Dr Nicholas M Redshaw PhD [99], Dr Stefanie V Lensing PhD [99], Shane McCarthy [99], Alex Makunin [99], Dr Carlos E Balcazar PhD [90], Dr Michael D Gallagher PhD [90], Dr Kathleen A Williamson PhD [90], Thomas D Stanton BSc [90], Ms Michelle L Michelsen BSc [91], Ms Joanna Warwick-Dugdale BSc [91], Dr Robin Manley Ph.D. [91], Ms Audrey Farbos MSc [91], Dr James W Harrison Ph.D. [91], Dr Christine M Sambles Ph.D. [91], Dr David J Studholme PhD. [91], Dr Angie Lackenby [66], Dr Tamyo Mbisa [66], Dr Steven Platt [66], Mr Shahjahan Miah [66], Dr David Bibby [66], Dr Carmen Manso [66], Dr Jonathan Hubb [66], Dr Gavin Dabrera [66], Dr Mary Ramsay [66], Dr Daniel Bradshaw [66], Dr Ulf Schaefer [66], Dr Natalie Groves [66], Dr Eileen Gallagher [66], Dr David Lee [66], Dr David Williams [66], Dr Nicholas Ellaby [66], Hassan Hartman [66], Nikos Manesis [66], Vineet Patel [66], Juan Ledesma [67], Ms Katherine A Twohig [67], Dr Elias Allara [64, 88], Ms Clare Pearson [64, 88], Mr Jeffrey K. J. Cheng MSc [94], Dr Hannah E. Bridgewater PhD [94], Ms Lucy R. Frost BSc [94], Ms Grace Taylor-Joyce BSc [94], Dr Paul E Brown PhD [94], Dr Lily Tong PhD

[48], Ms Alice Broos BSc [48], Mr Daniel Mair BSc [48], Mrs Jenna Nichols BSc [48], Dr Stephen N Carmichael PhD [48], Dr Katherine L Smollett PhD [40], Dr Kyriaki Nomikou PhD [48], Dr Elihu Aranday-Cortes PhD/DVM [48], Ms Natasha Johnson BSc [48], Dr Seema Nickbakhsh PhD [48, 68], Dr Edith E Vamos PhD [92], Dr Margaret Hughes PhD [92], Dr Lucille Rainbow PhD [92], Mr Richard Eccles MSc [92], Ms Charlotte Nelson MSc [92], Dr Mark Whitehead PhD [92], Dr Richard Gregory PhD [92], Mr Matthew Gemmell MSc [92], Ms Claudia Wierzbicki BSc [92], Ms Hermione J Webster BSc [92], Ms Chloe L Fisher MSc [28], Mr Adrian W Signell BSc [20], Dr Gilberto Betancor PhD [20], Mr Harry D Wilson BSc [20], Dr Gaia Nebbia PhD FRCPath [12], Dr Flavia Flaviani PhD [31], Mr Alberto C Cerda MSc [96], Ms Tammy V Merrill MSc [96], Rebekah E Wilson MSc [96], Mr Marius Cotic MSc [82], Miss Nadua Bayzid BSc [82], Dr Thomas Thompson PhD [72], Dr Erwan Acheson PhD [72], Prof Steven Rushton PhD [51], Prof Sarah O'Brien PhD [51], David J Baker BEng [70], Steven Rudder [70], Alp Aydin MSci [70], Dr Fei Sang PhD [18], Dr Johnny Debebe PhD [18], Dr Sarah Francois PhD [23], Dr Tetyana I Vasylyeva DPhil [23], Dr Marina Escalera Zamudio PhD [23], Mr Bernardo Gutierrez MSc [23], Dr Angela Marchbank BSc [10], Joshua Maksimovic FD [9], Karla Spellman FD [9], Kathryn McCluggage Msc [9], Dr Mari Morgan PhD [69], Robert Beer BSc [9], Safiah Afifi BSc [9], Trudy Workman HNC [10], William Fuller BSc [10], Catherine Bresner Bsc [10], Dr Adrienn Angyal PhD [93], Dr Luke R Green PhD [93], Dr Paul J Parsons PhD [93], Miss Rachel M Tucker MSc [93], Dr Rebecca Brown PhD [93] and Mr Max Whiteley PhD [93]

**Software and analysis tools:**

James Bonfield BSc [99], Dr Christoph Puethe [99], Mr Andrew Whitwham BSc [99], Jennifier Liddle [99], Dr Will Rowe PhD [41], Dr Igor Siveroni PhD [39], Dr Thanh Le-Viet PhD [70] and Amy Gaskin MSc [69].

**Visualisation:**

Dr Rob Johnson PhD [39].

**1** Barking, Havering and Redbridge University Hospitals NHS Trust, **2** Basingstoke Hospital, **3** Belfast Health & Social Care Trust, **4** Betsi Cadwaladr University Health Board, **5** Big Data Institute, Nuffield Department of Medicine, University of Oxford, **6** Brighton and Sussex University Hospitals NHS Trust, **7** Cambridge Stem Cell Institute, University of Cambridge, **8** Cambridge University Hospitals NHS Foundation Trust, **9** Cardiff and Vale University Health Board, **10** Cardiff University, **11** Centre for Clinical Infection & Diagnostics Research, St. Thomas' Hospital and Kings College London, **12** Centre for Clinical Infection and Diagnostics Research, Department of Infectious Diseases, Guy's and St Thomas' NHS Foundation Trust, **13** Centre for Enzyme Innovation, University of Portsmouth (PORT), **14** Centre for Genomic Pathogen Surveillance, University of Oxford, **15** Clinical Microbiology Department, Queens Medical Centre, **16** Clinical Microbiology, University Hospitals of Leicester NHS Trust, **17** County Durham and Darlington NHS Foundation Trust, **18** Deep Seq, School of Life Sciences, Queens Medical Centre, University of Nottingham, **19** Department of Infection Biology, Faculty of Infectious & Tropical Diseases, London School of Hygiene & Tropical Medicine, **20** Department of Infectious Diseases, King's College London, **21** Department of Microbiology, Kettering General Hospital, **22** Departments of Infectious Diseases and Microbiology, Cambridge University Hospitals NHS Foundation Trust; Cambridge, UK, **23** Department of Zoology, University of Oxford, **24** Division of Virology, Department of Pathology, University of Cambridge, **25** East Kent Hospitals University NHS Foundation Trust, **26** East Suffolk and North Essex NHS Foundation Trust, **27** Gateshead Health NHS Foundation Trust, **28** Genomics Innovation Unit, Guy's and St. Thomas' NHS Foundation Trust, **29** Gloucestershire Hospitals NHS Foundation Trust, **30** Great Ormond Street Hospital for Children NHS Foundation Trust, **31** Guy's and St. Thomas' BRC, **32** Guy's and St. Thomas' Hospitals, **33** Hampshire Hospitals NHS Foundation Trust, **34** Health Data Research UK Cambridge, **35** Health Services

Laboratories, **36** Heartlands Hospital, Birmingham, **37** Hub for Biotechnology in the Built Environment, Northumbria University, **38** Imperial College Hospitals NHS Trust, **39** Imperial College London, **40** Institute of Biodiversity, Animal Health & Comparative Medicine, **41** Institute of Microbiology and Infection, University of Birmingham, **42** King's College London, **43** Liverpool Clinical Laboratories, **44** Maidstone and Tunbridge Wells NHS Trust, **45** Manchester University NHS Foundation Trust, **46** Microbiology Department, Wye Valley NHS Trust, Hereford, **47** MRC Biostatistics Unit, University of Cambridge, **48** MRC-University of Glasgow Centre for Virus Research, **49** National Infection Service, PHE and Leeds Teaching Hospitals Trust, **50** Newcastle Hospitals NHS Foundation Trust, **51** Newcastle University, **52** NHS Greater Glasgow and Clyde, **53** NHS Lothian, **54** Norfolk and Norwich University Hospital, **55** Norfolk County Council, **56** North Cumbria Integrated Care NHS Foundation Trust, **57** North Tees and Hartlepool NHS Foundation Trust, **58** Northumbria University, **59** Oxford University Hospitals NHS Foundation Trust, **60** PathLinks, Northern Lincolnshire & Goole NHS Foundation Trust, **61** Portsmouth Hospitals University NHS Trust, **62** Princess Alexandra Hospital Microbiology Dept., **63** Public Health Agency, **64** Public Health England, **65** Public Health England, Clinical Microbiology and Public Health Laboratory, Cambridge, UK, **66** Public Health England, Colindale, **67** Public Health England, Colindale, **68** Public Health Scotland, **69** Public Health Wales NHS Trust, **70** Quadram Institute Bioscience, **71** Queen Elizabeth Hospital, **72** Queen's University Belfast, **73** Royal Devon and Exeter NHS Foundation Trust, **74** Royal Free NHS Trust, **75** Sandwell and West Birmingham NHS Trust, **76** School of Biological Sciences, University of Portsmouth (PORT), **77** School of Pharmacy and Biomedical Sciences, University of Portsmouth (PORT), **78** Sheffield Teaching Hospitals, **79** South Tees Hospitals NHS Foundation Trust, **80** Swansea University, **81** University Hospitals Southampton NHS Foundation Trust, **82** University College London, **83** University Hospital Southampton NHS Foundation Trust, **84** University Hospitals Coventry and Warwickshire, **85** University of Birmingham, **86** University of Birmingham Turnkey Laboratory, **87** University of Brighton, **88** University of Cambridge, **89** University of East Anglia, **90** University of Edinburgh, **91** University of Exeter, **92**

University of Liverpool, **93** University of Sheffield, **94** University of Warwick, **95** University of Cambridge, **96** Viapath, Guy's and St Thomas' NHS Foundation Trust, and King's College Hospital NHS Foundation Trust, **97** Virology, School of Life Sciences, Queens Medical Centre, University of Nottingham, **98** Wellcome Centre for Human Genetics, Nuffield Department of Medicine, University of Oxford, **99** Wellcome Sanger Institute, **100** West of Scotland Specialist Virology Centre, NHS Greater Glasgow and Clyde, **101** Department of Medicine, University of Cambridge, **102** Ministry of Health, Sri Lanka, **103** NIHR Health Protection Research Unit in HCAI and AMR, Imperial College London, **104** North West London Pathology, **105** NU-OMICS, Northumbria University, **106** University of Kent, **107** University of Oxford, **108** University of Southampton, **109** University of Southampton School of Health Sciences, **110** University of Southampton School of Medicine, **111** University of Surrey, **112** Warwick Medical School and Institute of Precision Diagnostics, Pathology, UHCW NHS Trust, **113** Wellcome Africa Health Research Institute Durban and **114** Wellcome Genome Campus.

**Supplementary File S4: Additional References**


W1 Green PJ. Reversible jump Markov chain Monte Carlo computation and Bayesian model determination. Biometrika. 1995; **82**: 711–32.

W2 Cauchemez S, Temime L, Guillemot D, *et al.* Investigating Heterogeneity in Pneumococcal Transmission. Journal of the American Statistical Association. 2006; **101**: 946–58.

W3 Meredith LW, Hamilton WL, Warne B, *et al.* Rapid implementation of SARS-CoV-2 sequencing to investigate cases of health-care associated COVID-19: a prospective genomic surveillance study. *Lancet Infect Dis* 2020; **20**: 1263–71.

W4 Contribution of nosocomial infections to the first wave in the United Kingdom. https://assets.publishing.service.gov.uk/government/uploads/system/uploads/attachment_data/file/961210/S1056_Contribution_of_nosocomial_infections_to_the_first_wave.pdf (accessed June 28, 2021).

W5 Challen R, Brooks-Pollock E, Tsaneva-Atanasova K, Danon L. Meta-analysis of the SARS-CoV-2 serial interval and the impact of parameter uncertainty on the COVID-19 reproduction number. DOI:10.1101/2020.11.17.20231548.

W6 Lauer SA, Grantz KH, Bi Q, *et al.* The Incubation Period of Coronavirus Disease 2019 (COVID-19) From Publicly Reported Confirmed Cases: Estimation and Application. *Ann Intern Med* 2020; **172**: 577–82.

W7 Boni MF, Lemey P, Jiang X, *et al.* Evolutionary origins of the SARS-CoV-2 sarbecovirus lineage responsible for the COVID-19 pandemic. *Nat Microbiol* 2020; **5**: 1408–17.
